# Using synthetic controls to estimate the population-level effects of Ontario’s recently implemented overdose prevention sites and consumption and treatment services

**DOI:** 10.1101/2021.12.13.21267739

**Authors:** Dimitra Panagiotoglou, Jihoon Lim

## Abstract

**Background:** Between 2017 and 2020, Ontario implemented overdose prevention sites (OPS) and consumption and treatment services (CTS) in nine of its 34 public health units (PHU). We tested for the effect of booth-hours (spaces within OPS/CTSs for supervised consumption) on opioid-related health service use and mortality rates at the provincial-(aggregate) and PHU-level.

**Methods:** We used monthly rates of all opioid-related emergency department (ED) visits, hospitalizations, and deaths between January 2015 and March 2021 as our three outcomes. For each PHU that implemented OPS/CTSs, we created a synthetic control as a weighted combination of unexposed PHUs. Our exposure was the time-varying rate of booth-hours provided. We estimated the population-level effects of the intervention on each outcome per treated/synthetic-control pair using controlled interrupted time series with segmented regression; and tested for the aggregate effect using a multiple baseline approach. We adjusted for time-varying provision of prescription opioids for pain management, opioid agonist treatment (OAT), and naloxone kits; and corrected for seasonality and autocorrelation. All rates were per 100,000 population. For sensitivity analysis, we restricted the post-implementation period to before COVID-19 public health measures were implemented (March 2020).

**Results:** Our aggregate analyses found no effect per booth-hour on ED visit (0.00, 95% CI: -0.01, 0.01; p-value=0.6684), hospitalization (0.00, 95% CI: 0.00, 0.00; p-value=0.9710) or deaths (0.00, 95% CI: 0.00, 0.00; p-value=0.2466). However, OAT reduced ED visits (−0.20, 95% CI: -0.35, -0.05; p-value=0.0103) and deaths (−0.04, 95% CI: -0.05, -0.03; p-value=<0.0001). Conversely, prescription opioids for pain management modestly increased deaths (0.0008, 95% CI: 0.0002, 0.0015; p-value=0.0157) per 100,000 population, respectively. Except for a few treated PHU/synthetic control pairs, disaggregate results were congruent with overall findings.

**Conclusion:** Booth-hours had no population-level effect on opioid-related overdose ED visit, hospitalization, or death rates.

## INTRODUCTION

Between January 2016 and June 2021, there were 24,626 opioid-related overdose deaths, and at least 27,604 opioid poisoning hospitalizations in Canada (Special Advisory Committee on the Epidemic of Opioid Overdoses, 2022). Although British Columbia is Canada’s hardest hit province, Ontario is a close second in opioid-related disease burden and health service use. Over a ten-year period (2007-2016, inclusive), Ontario’s emergency department (ED) visits increased 50% to 55.3 per 100,000 population (Tam, 2018) and hospitalizations rose 12% to 14.8 per 100,000 population (Canadian Institute for Health Information, 2018). For the first time in decades, male life expectancy at birth decreased as a consequence of high opioid-related mortality observed in younger adults (25 – 44 years of age) (Statistics Canada, 2020).

Overdose prevention sites (OPS) and consumption and treatment services (CTS) are among a suite of harm reduction interventions (e.g., supervised injection facilities, needle and syringe programs, naloxone distribution programs, drug checking services, and opioid agonist treatment (OAT)) increasingly offered across Canada to mitigate the negative physical and social consequences of illicit drug use (Strike & Watson, 2019). Historically unsanctioned, OPSs are temporary sites providing nimble, grassroots, peer-managed responses to the neglected needs of people who use illicit substances. Aside from providing critical overdose reversal services, OPSs offer overdose prevention education, Take Home Naloxone training and distribution, access to drug use equipment, and safe disposal of used equipment (BC Centre for Disease Control, 2019; Ministry of Health and Long-Term Care, 2018a). As of 2017, OPSs no longer require federal government (Health Canada) exemption under section 56.1 of the Controlled Drugs and Substances Act during public health emergencies. Conversely, CTSs (elsewhere known as supervised consumption sites) have Health Canada approval and must have a designated health professional (e.g., nurse) on-site. Aside from the harm reduction services available at OPSs, CTSs must also offer or provide a defined, proximal pathway to addictions treatment services (i.e., OAT, detox, residential or community treatment), primary care, mental health, and housing or social assistance programs (Ministry of Health and Long-Term Care, 2018a). Notably, CTSs do not offer supervised inhalation services.

While some authors have concluded that supervised consumption sites (namely, safe injection facilities) reduce mortality and health service use, they remain politically controversial (Potier C, Laprévote V, Dubois-Arber F, Cottencin O, & Rolland B, 2014). Much of the available evidence is from the concentrated drug use epidemics of Vancouver’s Downtown Eastside and Sydney’s ‘red light’ district and specific to injection drug use (Kennedy, Hayashi, Milloy, Wood, & Kerr, 2019; Potier C et al., 2014). The effects of supervised consumption site variants such as OPSs and CTSs remain unclear, particularly in contexts where the population is geographically diffuse, services are not restricted to people who inject drugs, and mobile rather than fixed sites operate. Further, most studies fail to distinguish the effects of supervised consumption sites from other harm reduction interventions also available (Caulkins, Pardo, & Kilmer, 2019). Our aim is to estimate the population-level effects of Ontario’s OPS/CTSs implemented between 2017 and 2020 (inclusive) on opioid-related ED visit, hospitalization, and death rates, while controlling for other time-varying interventions including rates of prescription opioids for pain management and opioid agonist treatment (separately) and naloxone kits distributed. Based on results from a similar analysis we conducted using the British Columbia Centre for Disease Control’s Overdose Cohort data, we hypothesize that OPS/CTSs had no effects on health service use and death rates once we control for co-occurring time-varying interventions (Panagiotoglou, 2022). Results from this study are pertinent to other communities with population demographics and needs different from Vancouver’s well studied Downtown Eastside.

## METHODS

### Study Design

We conducted a retrospective, ecological study that uses variations in the provision of OPS/CTS across Ontario’s public health units (PHU) to examine population-level health outcomes.

### Setting

Ontario is Canada’s most populous province with approximately 14.75 million residents (Government of Ontario, 2020), 97% of whom are covered by the provincial health insurance program (Izenberg, Iroanyah, & Thompson, 2018). Within the province, health promotion and disease prevention are administered by PHUs, which have mutually exclusive and exhaustive health boundaries. The 34 PHUs range from 33,166 residents in Timiskaming, the province’s most sparsely populated health unit, to 3,094,237 for its densest, most populated urban unit, Toronto (Government of Ontario, 2020).

Beginning in 2016, several provincial and federal harm reduction interventions were implemented to stem the rising rate of opioid-related overdose deaths. In June 2016, the Ontario Naloxone Program for Pharmacies (ONPP) began offering naloxone injection kits at no charge to individuals at risk of opioid-related overdose or persons in a position to assist someone at risk of an overdose, while select community-based organizations continued to dispense naloxone to at-risk clients, friends and family through a program that first began in 2013 (Canadian Centre on Substance Abuse & Canadian Community Epidemiology Network on Drug Use, 2016; Canadian Pharmacists Association, 2017). Shortly thereafter, naloxone kits transitioned from an injectable formulation to nasal spray (January 2017), the federal government’s Good Samaritan Drug Overdose Act became law (May 2017) (Government of Canada, 2018), and PHUs began sub-distributing naloxone kits to community-based organizations much more widely through the Ontario Naloxone Program (ONP, September 2017) (Gogolishvili & Wasdell, 2020). Although some cities began operating unsanctioned sites earlier in the year, Health Canada issued an exemption to Ontario to establish legally sanctioned temporary OPSs on December 7, 2017 (Ontario Ministry of Health & Ontario Ministry of Long-Term Care, 2018). Beginning October 2018, the government of Ontario enforced a list of required services and metrics that needed to be met for continued approval and financial support of CTSs (Ministry of Health and Long-Term Care, 2018a) and applied a cap to the number of sites allowed, ostensibly restricting provision of sites to larger cities (Russell, Imtiaz, Ali, Elton-Marshall, & Rehm, 2020).

Between June 2017 and December 2020, nine of the 34 provincial PHUs began operating at least one OPS/CTSs. These were in Toronto (n=9, 2 additional shelter-hotel sites), Ottawa (n=5), and one each in Hamilton, Kingston, London, Niagara, Thunder Bay, Guelph, and Waterloo (Pivot Legal Society, 2020) (Supplementary Table 1 for implementation dates, hours of operation, and capacity).

We excluded two PHUs that merged during the study period (Southwestern and Huron-Perth) with a combined population of approximately 336,000 (2.3%) from subsequent analysis owing to the incomplete demographic data available and differences between public health units’ and Statistics Canada’s jurisdictional boundaries.

### Data collection and measures

We used monthly counts of all opioid-related ED visits, acute care hospitalizations, and deaths between January 2014 and March 2021, reported at the PHU-level in Ontario’s publicly accessible Interactive Opioid Tool (Ontario Agency for Health Protection and Promotion (Public Health Ontario), 2020). Across the 32 PHUs included in the study, there were 53,806 ED visits, 13,734 hospitalizations and 9,377 mortalities for opioid-related overdose events during this time.

#### Opioid overdose related emergency department visits, acute care hospitalizations, and deaths

The Interactive Opioid Tool includes all opioid-related ED visits and acute care hospitalizations as captured in the province-wide National Ambulatory Care Reporting System (NACRS) and Discharge Abstract Database (DAD), respectively. We included all fatal events where accidental opioid poisoning from codeine, fentanyl (including carfentanil and other fentanyl analogues), heroin, hydrocodone, hydromorphone, methadone, morphine, or oxycodone was considered a contributing cause of death according to the Office of the Chief Coroner are also included. More than one opioid can be present at time of death, and presence of a drug does not necessarily indicate that it contributed to death. We converted event counts to rates per 100,000 to enable comparability between PHUs using the Ontario Ministry of Health and Long-Term Care’s IntelliHealth population level estimates (2003-2016) and population projections (2017 – 2021) also available on the Interactive Opioid Tool.

#### Other data

We supplemented the PHU-level overdose event data with population-level annual demographics (percent of the population without a high school diploma, immigrant, and visible minority; median household income); age- and sex-standardized rates of alcohol-related emergency department visits and hospitalizations; and monthly rates per 100,000 of concomitant interventions: persons receiving opioid prescriptions (for pain management and OAT, separately), and naloxone kits distributed. Finally, we created a measure of OPS/CTS intervention ‘intensity’ as the sum of the product of individual booth-spaces and hours available for supervised consumption of illicit substances across sites and standardized it to booth-hours per 100,000 population. For full definitions, data sources, and methods, please see supplementary material.

### Ethics approval

This study was exempt from ethics review by McGill University’s Institutional Review Board.

### Statistical analysis

We used controlled interrupted time series with segmented regression to test the effects of OPS/CTS booth-hours on opioid-related ED visits, hospitalizations, or deaths (Bernal JL, Cummins S, & Gasparrini A, 2017). Interrupted time series analysis is a quasi-experimental study design that estimates population-level effects of health service or policy interventions before and after implementation in contexts where randomized controlled trials are not feasible or ethical (Cook TD & DT, 1979; Zhang F, Wagner AK, & Ross-Degnan D, 2011). The advantage of including a well-chosen control as the counterfactual is that selection bias and within-group characteristics that change slowly over time, secular changes, random fluctuations from one time point to the next and regression to the mean are also controlled (Lopez Bernal, Cummins, & Gasparrini, 2018).

Treated groups included all PHUs that implemented at least one OPS/CTS during the study period. Synthetic controls were derived for each treated PHU per outcome from a donor pool of exclusively never-treated PHUs (Baker, Larcker, & Wang, 2021) using Abadie et al.’s data-driven method and the Synth package with its extensions in R (Abadie, Diamond, & Hainmueller, 2011; Castanho Silva & DeWitt, 2020; Degli Esposti et al., 2020). The pre-intervention period was restricted to two years to optimize prediction of the treatment group by the comparison group (Bilinski, 2021).

We fitted the following linear regression model for intervention status *j* (pre-intervention=0, post-intervention=1), for group *k* (control group=0, treated group=1), at time *t* (pre-intervention implementation <0, month of intervention implementation=0, post-intervention implementation>0):

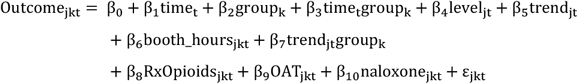

Where β_0_ is the outcome rate intercept for the control; β_1_ represents the pre-intervention temporal trend for the control’s outcome rate; β_2_ indicates the pre-intervention difference between the treated group’s outcome rate intercept and the controls (level difference); β_3_ captures the pre-intervention difference between treated and control groups’ temporal trends; β_4_ represents the difference in the control outcome rate immediately post-intervention compared with the control outcome rate at the beginning of the pre-intervention observation period (level difference); β_5_ indicates the change in temporal trend for the control outcome rate post-intervention compared with pre-intervention; β_6_ is the effect of the booth-hours on the treated group’s outcome rate; and β_7_ indicates the difference between treated and control groups’ temporal trends post-implementation (i.e., the treated groups change in trend *relative* to the control group’s change in trend, β_5_). We also adjusted for other relevant time-varying confounders following the disjunctive cause criterion for confounder selection such that all covariates are known or potential causes of the exposure (i.e., OPS/CTS implementation) and/or the outcome (i.e., opioid-related ED visits, hospitalizations or death) (VanderWeele & Shpitser, 2011). Finally, we corrected for seasonality using harmonic terms, and used Newey-West standard errors to account for autocorrelation identified using plots of residuals.

Since the initial OPS/CTSs was implemented at different times across the nine PHUs, we applied a multiple baseline approach (i.e. using intervention time instead of calendar time)(Hawkins, Sanson-Fisher, Shakeshaft, D’Este, & Green, 2007) to measure the effect of booth-hours on population-level ED visits, hospitalizations, and deaths. We aggregated the data across treated PHUs with well-fitted synthetic controls and report the results of these aggregate analyses and their sensitivity analyses in the main text. We also report the disaggregated results per PHU treated/synthetic control pair in the supplementary material.

For sensitivity analysis, we tested the effects of the Good Samaritan Drug Overdose Act(Government of Canada, 2018) across all treated/synthetic control pairs (not shown here), aggregated all treated PHU/synthetic control pairs irrespective of fit, and terminated the observation period to before March 2020 and the onset of COVID-19 related service restrictions and border closures.

All analyses were conducted using R version 4.0.5(R Core Team, 2014). All data used for this study are publicly available through the Ontario Opioid Tool, Ontario Drug Policy Research Network, news articles and OPS/CTS listed hours of operation (see Supplement). All results are reported using 95% CI and accompanying p-values. Because we created a unique synthetic control per treated PHU *per* outcome, we did not use a p-value correction for interpreting our results. However, a simple Bonferroni correction per treated unit assuming we were using the *same* synthetic control across all three outcomes would change the accepted p-value from 0.05 to 0.017.

## RESULTS

When the first unsanctioned OPSs began operating in Toronto and Ottawa, Hamilton (n=9, 1.58 per 100,000), Niagara Region (n=7, 1.53 per 100,000), Toronto (n=27, 0.92 per 100,000), Peel (n=11, 0.73 per 100,000), Ottawa (n=7, 0.70 per 100,000), and York (n=7, 0.59 per 100,000) reported the highest opioid-related mortality counts; while Kingston-Frontenac-Lennox-Addison (n=5, 2.42 per 100,000), North Bay-Parry Sound (n=3, 2.34 per 100,000), Peterborough (n=3, 2.11 per 100,000), Thunder Bay (n=3, 1.95 per 100,000), and Haldimand-Norfolk (n=2, 1.79 per 100,000) had the highest mortality rates. Meanwhile, small, less urban PHUs (Chatham-Kent, Eastern Ontario, Haliburton-Kawartha-Pine Ridge, Hastings-Prince Edward, Leeds-Grenville, Porcupine, Renfrew, Timiskaming, and Wellington-Dufferin-Guelph) reported no mortality events. These patterns persisted for ED and hospitalization events such that the largest, most urban PHUs did not have the highest event rates despite high event counts. Except for Niagara and Thunder Bay, treated and potential control units in the donor pool were sufficiently similar in population demographics and outcomes (i.e. none of the treated units were outside the convex hull of the donor pool), overdose risks and relevant policies; with no known external shocks specific to control units (Bouttell, Craig, Lewsey, Robinson, & Popham, 2018). Niagara and Thunder Bay’s outcomes were consistently outside the convex hull in the six months prior to intervention implementation such that suitable synthetic controls could not be established across any of the outcomes. The same occurred for London ED visits and hospitalizations, and Kingston hospitalizations. In these cases, we excluded the poorly matched treated PHU/synthetic control pairs from the primary aggregate analyses but report results where they are included as well (see Table 4).

Tables 1 and 2 show the predictor weights and public health units used to create the synthetic controls per treated unit by outcome.

**Table 1.**
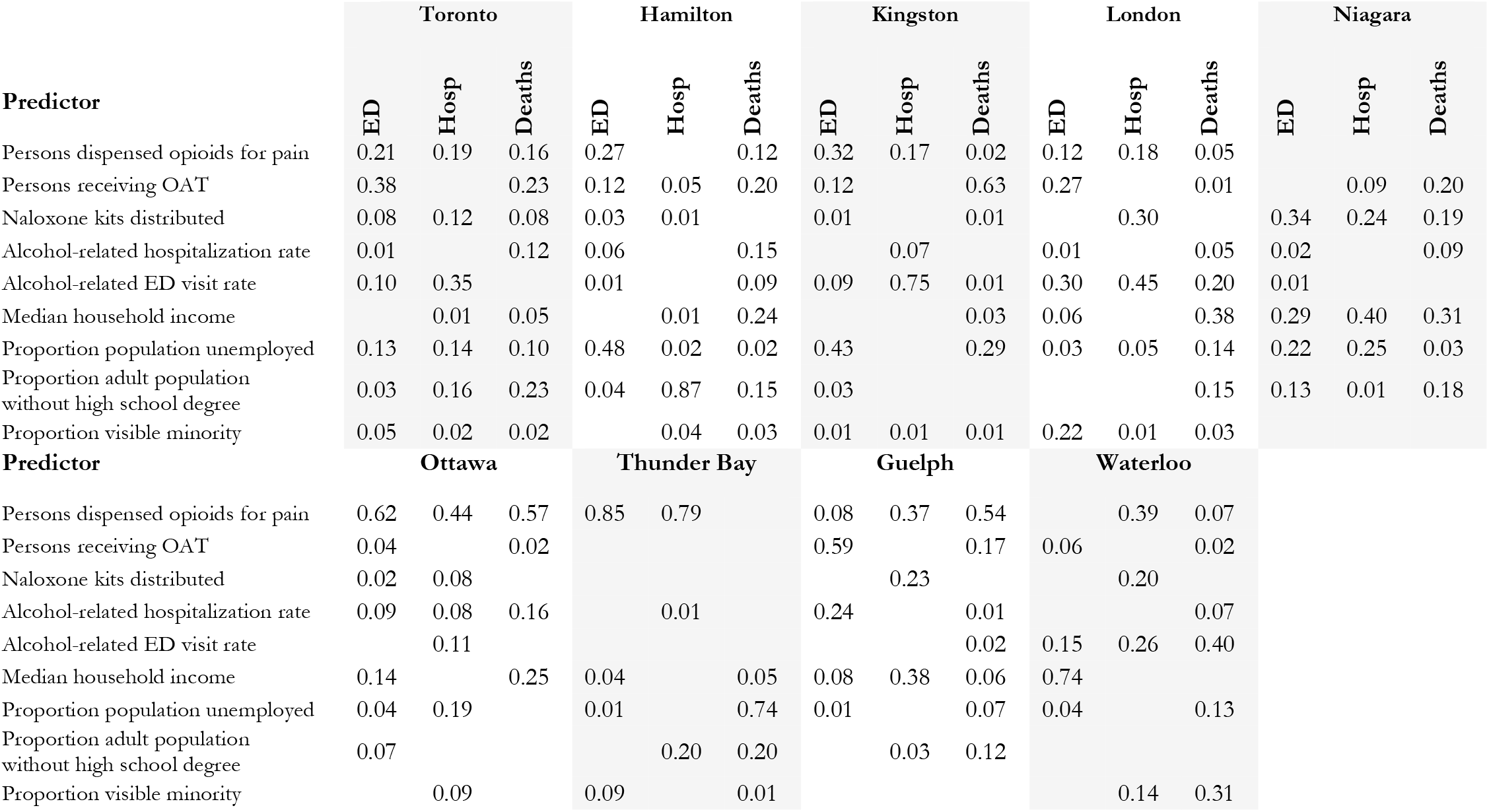
Synthetic control predictor weights ED=Emergency department; Hosp=Hospitalization; OAT=Opioid agonist treatment; pop=population

**Table 2.** Synthetic control donor public health units

| Public Health Unit | Toronto |  |  | Hamilton |  |  | Kingston |  |  | London |  |  |
| --- | --- | --- | --- | --- | --- | --- | --- | --- | --- | --- | --- | --- |
|  | ED | Hosp | Deaths | ED | Hosp | Deaths | ED | Hosp | Deaths | ED | Hosp | Deaths |
| Algoma |  |  | 0.09 |  |  |  |  |  |  |  |  |  |
| Brant County |  |  |  | 0.19 | 0.15 | 0.30 | 0.26 | 0.17 | 0.22 | 0.08 |  |  |
| Chatham-Kent |  |  |  |  |  |  |  |  |  |  | 0.20 | 0.09 |
| Durham Region |  | 0.18 |  |  |  |  |  |  |  |  |  |  |
| Eastern Ontario |  |  |  |  |  |  |  |  |  |  | 0.26 |  |
| Grey Bruce |  |  |  | 0.04 |  |  |  |  | 0.24 |  |  |  |
| Haldimand-Norfolk |  |  |  | 0.28 |  | 0.07 |  |  |  |  |  |  |
| Haliburton-Kawartha-Pine Ridge |  |  |  |  |  |  |  |  |  |  |  |  |
| Halton Region |  |  |  |  |  |  | 0.22 | 0.07 | 0.01 |  |  |  |
| Hastings Prince Edward |  |  |  |  |  |  |  |  |  |  |  |  |
| Lambton |  |  |  |  |  |  |  |  |  |  | 0.16 |  |
| Leeds-Grenville-Lark |  |  | 0.06 |  |  |  |  | 0.08 |  |  |  | 0.54 |
| North Bay Parry Sound |  |  |  |  |  |  |  |  |  |  |  |  |
| Northwestern | 0.24 |  | 0.01 | 0.03 | 0.13 | 0.02 |  | 0.03 | 0.14 | 0.27 | 0.18 |  |
| Peel | 0.40 | 0.67 | 0.50 |  | 0.07 |  |  |  |  | 0.16 | 0.19 |  |
| Peterborough |  |  |  | 0.13 | 0.17 | 0.26 | 0.30 | 0.19 | 0.15 | 0.12 | 0.01 | 0.14 |
| Porcupine |  |  |  |  |  |  |  |  |  |  |  |  |
| Renfrew |  |  |  |  |  |  |  |  |  |  |  |  |
| Simcoe Muskoka | 0.21 |  | 0.34 | 0.34 | 0.48 |  |  |  | 0.22 |  |  |  |
| Sudbury |  | 0.03 |  |  |  | 0.35 | 0.22 | 0.30 |  | 0.36 |  |  |
| Timiskaming |  |  |  |  |  |  |  |  |  |  |  |  |
| Windsor-Essex |  |  |  |  |  |  |  |  |  |  |  | 0.23 |
| York | 0.15 | 0.12 |  |  |  |  | 0 | 0.15 |  |  |  |  |
| PHU with non-zero weights | 4 | 4 | 5 | 6 | 5 | 5 | 4 | 7 | 6 | 5 | 6 | 4 |

| Public Health Unit | Niagara |  |  | Ottawa |  |  | Thunder Bay |  |  | Guelph |  |  | Waterloo |  |  |
| --- | --- | --- | --- | --- | --- | --- | --- | --- | --- | --- | --- | --- | --- | --- | --- |
|  | ED | Hosp | Deaths | ED | Hosp | Deaths | ED | Hosp | Deaths | ED | Hosp | Deaths | ED | Hosp | Deaths |
| Algoma |  |  |  |  |  |  |  |  |  |  |  |  |  |  |  |
| Brant County | 0.13 | 0.13 | 0.26 |  |  |  | 0.20 | 0.01 | 0.38 | 0.07 |  | 0.28 | 0.34 | 0.01 | 0.19 |
| Chatham-Kent |  |  |  |  |  |  |  |  |  |  |  |  |  | 0.01 |  |
| Durham Region |  |  | 0.01 |  | 0.18 |  |  |  |  |  |  |  |  | 0.01 |  |
| Eastern Ontario |  | 0.01 | 0.01 |  | 0.08 |  |  |  |  |  | 0.18 |  |  | 0.34 |  |
| Grey Bruce | 0.01 | 0.01 | 0.01 |  |  |  |  |  |  | 0.19 | 0.24 | 0.27 | 0.27 | 0.01 | 0.22 |
| Haldimand-Norfolk |  |  | 0.01 |  |  |  |  |  |  |  |  |  |  | 0.01 |  |
| Haliburton-Kawartha-Pine Ridge |  | 0.01 | 0.01 |  |  |  |  |  |  |  |  |  |  | 0.01 |  |
| Halton Region |  |  | 0.03 | 0.10 | 0.27 |  |  |  |  | 0.25 | 0.33 | 0.12 | 0.26 | 0.37 | 0.34 |
| Hastings Prince Edward | 0.17 | 0.24 | 0.01 |  |  |  |  |  |  |  |  |  |  | 0.01 |  |
| Lambton |  | 0.03 | 0.24 |  |  |  |  |  |  |  |  |  |  | 0.01 |  |
| Leeds-Grenville-Lark | 0.01 | 0.01 | 0.01 | 0.02 |  |  |  |  |  |  |  |  |  | 0.01 |  |
| North Bay Parry Sound |  |  |  |  |  |  | 0.26 | 0.34 |  |  |  |  |  |  |  |
| Northwestern |  |  | 0.01 | 0.01 | 0.07 | 0.05 | 0.08 | 0.39 | 0.35 | 0.04 |  |  |  | 0.03 |  |
| Peel |  |  | 0.03 | 0.21 | 0.02 | 0.35 |  |  |  |  |  |  |  |  |  |
| Peterborough | 0.58 | 0.54 | 0.25 |  |  |  |  | 0.20 | 0.09 |  |  |  |  |  |  |
| Porcupine |  |  |  |  |  |  |  |  |  |  | 0.09 |  |  | 0.01 |  |
| Renfrew |  | 0.01 | 0.01 |  |  |  |  |  |  |  |  |  |  | 0.01 |  |
| Simcoe Muskoka |  |  | 0.01 | 0.25 | 0 | 0.21 |  |  |  | 0.29 |  | 0.01 |  |  |  |
| Sudbury |  |  | 0.05 |  |  |  | 0.47 |  | 0.18 |  |  |  | 0.12 | 0.01 | 0.09 |
| Timiskaming |  |  |  |  |  |  |  | 0.01 |  |  |  |  |  |  |  |
| Windsor-Essex | 0.08 | 0.01 | 0.03 |  |  |  |  | 0.03 |  |  |  |  |  |  |  |
| York |  |  | 0.02 | 0.41 | 0.38 | 0.39 |  |  |  | 0.17 | 0.14 | 0.32 |  | 0.15 | 0.16 |
| PHU with non-zero weights | 6 | 10 | 18 | 6 | 6 | 4 | 4 | 6 | 4 | 6 | 5 | 5 | 4 | 16 | 5 |

Table 3 shows the balance between treated and synthetic controls’ predictors compared with the overall sample means at time of intervention implementation, per outcome. Notably, perfect balance is not observed across all predictors for each treated-synthetic control unit given the weight of each predictor per outcome ranged from 0.00 (no effect in creating synthetic control for that outcome) to 0.85 for persons prescribed opioids for pain management when creating Thunder Bay’s synthetic control for ED visits (treated: 4412, synthetic control: 4408, sample mean: 3838).

**Table 3.**
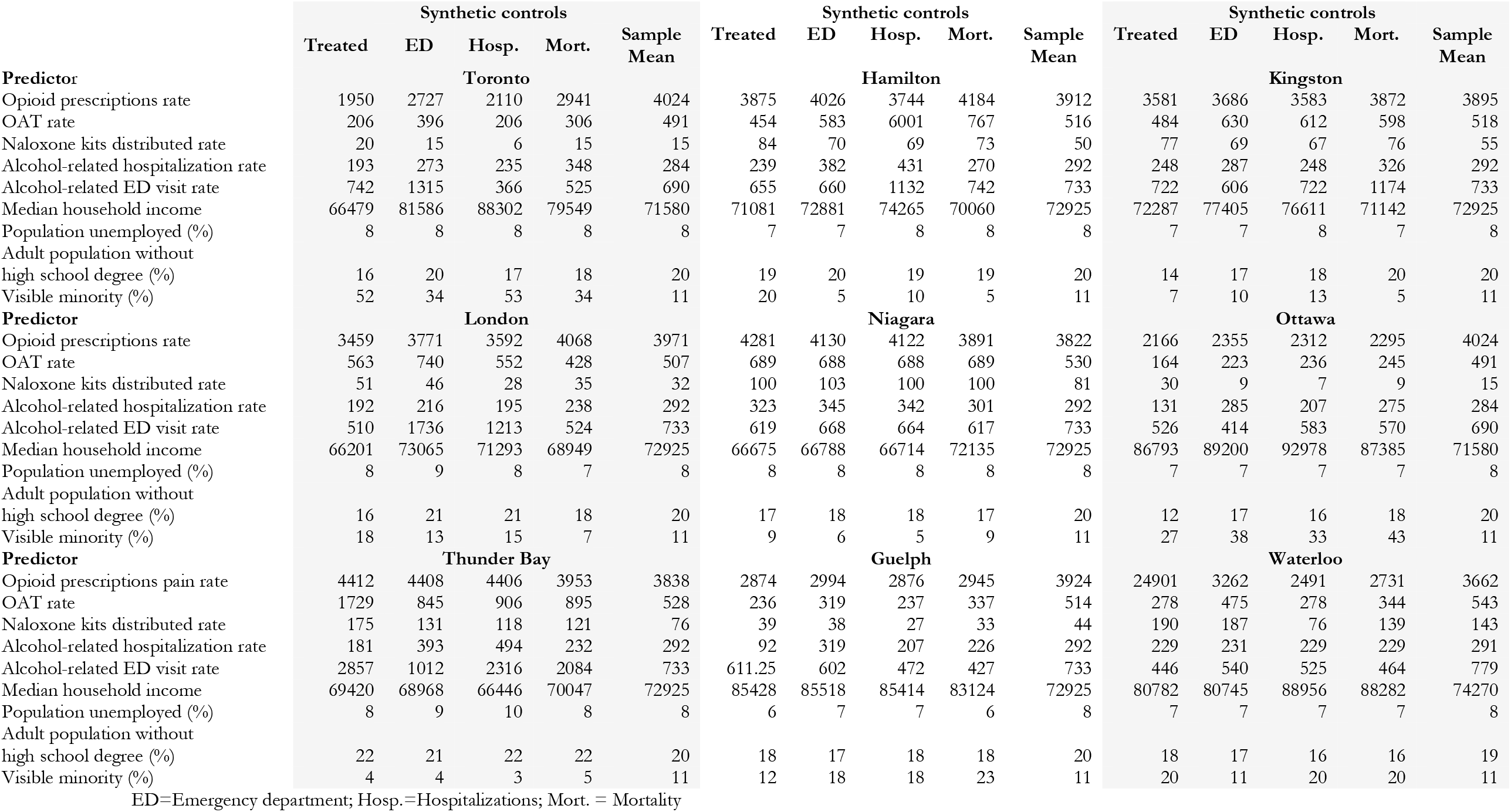
Characteristics of treated units, synthetic controls (per outcome) and overall sample mean at time of intervention; rate per 100,000 population ED=Emergency department; Hosp.=Hospitalizations; pop.=population; Mort.=Mortalities

| Predictor | Synthetic controls |  |  |  |  | Synthetic controls |  |  |  |  | Synthetic controls |  |  |  |  |
| --- | --- | --- | --- | --- | --- | --- | --- | --- | --- | --- | --- | --- | --- | --- | --- |
|  | Treated | ED | Hosp. | Mort. | Sample Mean | Treated | ED | Hosp. | Mort. | Sample Mean | Treated | ED | Hosp. | Mort. | Sample Mean |
|  | Toronto |  |  |  |  | Hamilton |  |  |  |  | Kingston |  |  |  |  |
| Opioid prescriptions rate | 1950 | 2727 | 2110 | 2941 | 4024 | 3875 | 4026 | 3744 | 4184 | 3912 | 3581 | 3686 | 3583 | 3872 | 3895 |
| OAT rate | 206 | 396 | 206 | 306 | 491 | 454 | 583 | 6001 | 767 | 516 | 484 | 630 | 612 | 598 | 518 |
| Naloxone kits distributed rate | 20 | 15 | 6 | 15 | 15 | 84 | 70 | 69 | 73 | 50 | 77 | 69 | 67 | 76 | 55 |
| Alcohol-related hospitalization rate | 193 | 273 | 235 | 348 | 284 | 239 | 382 | 431 | 270 | 292 | 248 | 287 | 248 | 326 | 292 |
| Alcohol-related ED visit rate | 742 | 1315 | 366 | 525 | 690 | 655 | 660 | 1132 | 742 | 733 | 722 | 606 | 722 | 1174 | 733 |
| Median household income | 66479 | 81586 | 88302 | 79549 | 71580 | 71081 | 72881 | 74265 | 70060 | 72925 | 72287 | 77405 | 76611 | 71142 | 72925 |
| Population unemployed (%) | 8 | 8 | 8 | 8 | 8 | 7 | 7 | 8 | 8 | 8 | 7 | 7 | 8 | 7 | 8 |
| Adult population without high school degree (%) | 16 | 20 | 17 | 18 | 20 | 19 | 20 | 19 | 19 | 20 | 14 | 17 | 18 | 20 | 20 |
| Visible minority (%) | 52 | 34 | 53 | 34 | 11 | 20 | 5 | 10 | 5 | 11 | 7 | 10 | 13 | 5 | 11 |
|  | London |  |  |  |  | Niagara |  |  |  |  | Ottawa |  |  |  |  |
| Opioid prescriptions rate | 3459 | 3771 | 3592 | 4068 | 3971 | 4281 | 4130 | 4122 | 3891 | 3822 | 2166 | 2355 | 2312 | 2295 | 4024 |
| OAT rate | 563 | 740 | 552 | 428 | 507 | 689 | 688 | 688 | 689 | 530 | 164 | 223 | 236 | 245 | 491 |
| Naloxone kits distributed rate | 51 | 46 | 28 | 35 | 32 | 100 | 103 | 100 | 100 | 81 | 30 | 9 | 7 | 9 | 15 |
| Alcohol-related hospitalization rate | 192 | 216 | 195 | 238 | 292 | 323 | 345 | 342 | 301 | 292 | 131 | 285 | 207 | 275 | 284 |
| Alcohol-related ED visit rate | 510 | 1736 | 1213 | 524 | 733 | 619 | 668 | 664 | 617 | 733 | 526 | 414 | 583 | 570 | 690 |
| Median household income | 66201 | 73065 | 71293 | 68949 | 72925 | 66675 | 66788 | 66714 | 72135 | 72925 | 86793 | 89200 | 92978 | 87385 | 71580 |
| Population unemployed (%) | 8 | 9 | 8 | 7 | 8 | 8 | 8 | 8 | 8 | 8 | 7 | 7 | 7 | 7 | 8 |
| Adult population without high school degree (%) | 16 | 21 | 21 | 18 | 20 | 17 | 18 | 18 | 17 | 20 | 12 | 17 | 16 | 18 | 20 |
| Visible minority (%) | 18 | 13 | 15 | 7 | 11 | 9 | 6 | 5 | 9 | 11 | 27 | 38 | 33 | 43 | 11 |
|  | Thunder Bay |  |  |  |  | Guelph |  |  |  |  | Waterloo |  |  |  |  |
| Opioid prescriptions pain rate | 4412 | 4408 | 4406 | 3953 | 3838 | 2874 | 2994 | 2876 | 2945 | 3924 | 24901 | 3262 | 2491 | 2731 | 3662 |
| OAT rate | 1729 | 845 | 906 | 895 | 528 | 236 | 319 | 237 | 337 | 514 | 278 | 475 | 278 | 344 | 543 |
| Naloxone kits distributed rate | 175 | 131 | 118 | 121 | 76 | 39 | 38 | 27 | 33 | 44 | 190 | 187 | 76 | 139 | 143 |
| Alcohol-related hospitalization rate | 181 | 393 | 494 | 232 | 292 | 92 | 319 | 207 | 226 | 292 | 229 | 231 | 229 | 229 | 291 |
| Alcohol-related ED visit rate | 2857 | 1012 | 2316 | 2084 | 733 | 611.25 | 602 | 472 | 427 | 733 | 446 | 540 | 525 | 464 | 779 |
| Median household income | 69420 | 68968 | 66446 | 70047 | 72925 | 85428 | 85518 | 85414 | 83124 | 72925 | 80782 | 80745 | 88956 | 88282 | 74270 |
| Population unemployed (%) | 8 | 9 | 10 | 8 | 8 | 6 | 7 | 7 | 6 | 8 | 7 | 7 | 7 | 7 | 8 |
| Adult population without high school degree (%) | 22 | 21 | 22 | 22 | 20 | 18 | 17 | 18 | 18 | 20 | 18 | 17 | 16 | 16 | 19 |
| Visible minority (%) | 4 | 4 | 3 | 5 | 11 | 12 | 18 | 18 | 23 | 11 | 20 | 11 | 20 | 20 | 11 |

ED=Emergency department; Hosp.=Hospitalizations; Mort. = Mortality

Although there is no consensus on a root mean square prediction error (RMSPE) threshold distinguishing well-matched from inadequately constructed synthetic controls, using the RMSPE and outputs from the adjusted segmented regression (β_3_), we found the monthly pre-intervention outcome trends were parallel for well-matched treated/synthetic control PHU pairs (Table 4, Supplementary Table 3). As expected, differences in pre-intervention trends were observed for London ED visits (0.17, 95% CI: 0.03, 0.30; p-value=0.0169) and hospitalizations (0.05, 95% CI: 0.02, 0.08; p-value=0.0027); Kingston hospitalizations (0.10, 95% CI: 0.06, 0.15; p-value<0.0001); Thunder Bay ED visits (0.23, 95% CI: 0.08, 0.38; p-value=0.0031) and deaths (0.08, 95% CI: 0.01, 0.15; p-value=0.0207). Trend differences were also observed for Hamilton (0.04, 95% CI: 0.01, 0.07; p-value=0.0180) and Waterloo deaths (0.05, 95% CI: 0.02, 0.08; p-value=0.0053) compared with their respective synthetic controls; but not for Niagara and its synthetic control pair. In our sensitivity analyses where we restricted analysis to observations prior to COVID-19 related public health measures (i.e., observation period ending Feb 2020), differences between treated and synthetic control trends persisted for Kingston hospitalizations (0.10, 95% CI: 0.05, 0.16; p-value=0.0005), and Thunder Bay ED visits (0.24, 95% CI: 0.06, 0.42; p-value=0.0126) and deaths (0.07, 95% CI: 0.02, 0.13).

**Table 4.**
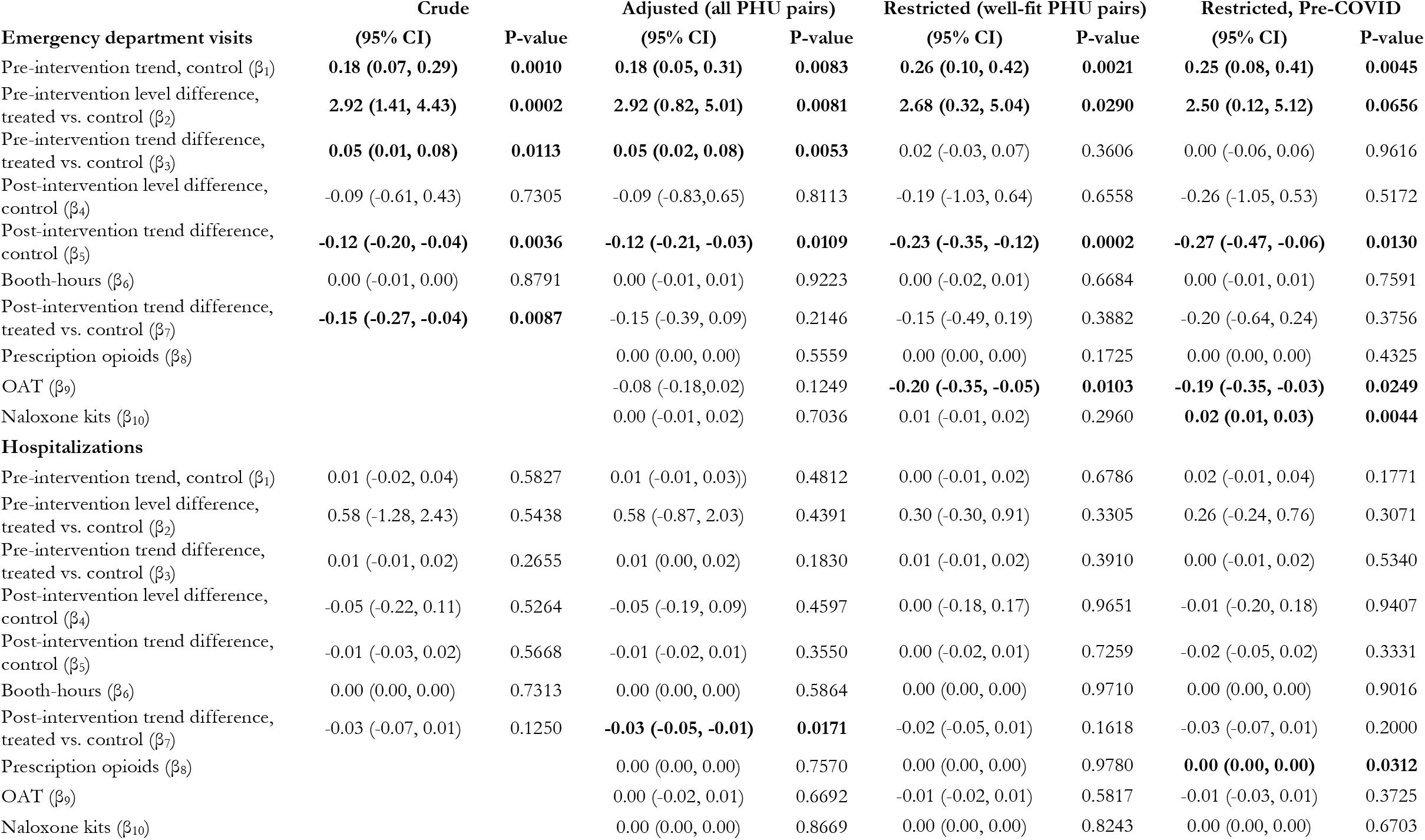
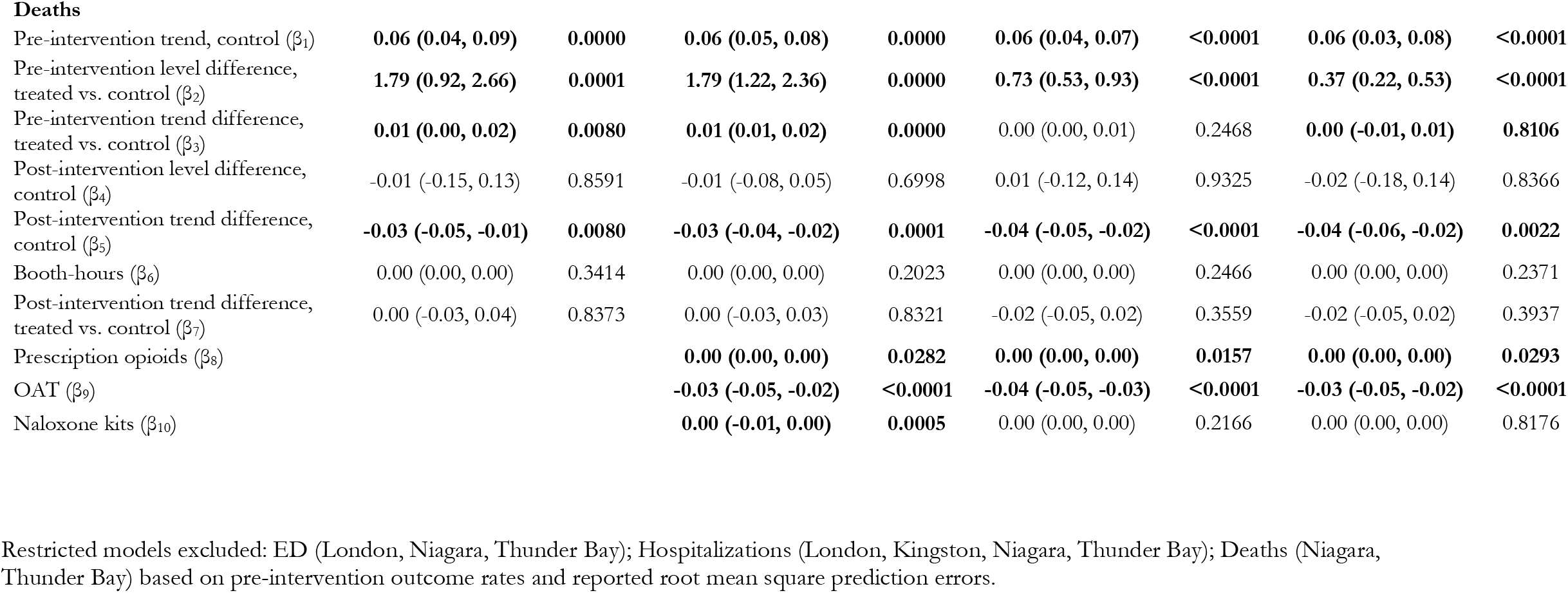
Aggregate analysis: crude, adjusted, restricted to PHUs with well-fitting synthetic controls, and analysis ending the post-intervention observation period on Feb 2020 (also restricted to well-fitted synthetic controls) *Adjusted with robust standard errors generated using Newey-West method

|  | Crude |  | Adjusted (all PHU pairs) |  | Restricted (well-fit PHU pairs) |  | Restricted, Pre-COVID |  |
| --- | --- | --- | --- | --- | --- | --- | --- | --- |
| Emergency department visits | (95% CI) | P-value | (95% CI) | P-value | (95% CI) | P-value | (95% CI) | P-value |
| Pre-intervention trend, control ($\beta_1$) | **0.18 (0.07, 0.29)** | **0.0010** | **0.18 (0.05, 0.31)** | **0.0083** | **0.26 (0.10, 0.42)** | **0.0021** | **0.25 (0.08, 0.41)** | **0.0045** |
| Pre-intervention level difference, treated vs. control ($\beta_2$) | **2.92 (1.41, 4.43)** | **0.0002** | **2.92 (0.82, 5.01)** | **0.0081** | **2.68 (0.32, 5.04)** | **0.0290** | **2.50 (0.12, 5.12)** | **0.0656** |
| Pre-intervention trend difference, treated vs. control ($\beta_3$) | **0.05 (0.01, 0.08)** | **0.0113** | **0.05 (0.02, 0.08)** | **0.0053** | 0.02 (-0.03, 0.07) | 0.3606 | 0.00 (-0.06, 0.06) | 0.9616 |
| Post-intervention level difference, control ($\beta_4$) | -0.09 (-0.61, 0.43) | 0.7305 | -0.09 (-0.83, 0.65) | 0.8113 | -0.19 (-1.03, 0.64) | 0.6558 | -0.26 (-1.05, 0.53) | 0.5172 |
| Post-intervention trend difference, control ($\beta_5$) | **-0.12 (-0.20, -0.04)** | **0.0036** | **-0.12 (-0.21, -0.03)** | **0.0109** | **-0.23 (-0.35, -0.12)** | **0.0002** | **-0.27 (-0.47, -0.06)** | **0.0130** |
| Booth-hours ($\beta_6$) | 0.00 (-0.01, 0.00) | 0.8791 | 0.00 (-0.01, 0.01) | 0.9223 | 0.00 (-0.02, 0.01) | 0.6684 | 0.00 (-0.01, 0.01) | 0.7591 |
| Post-intervention trend difference, treated vs. control ($\beta_7$) | **-0.15 (-0.27, -0.04)** | **0.0087** | -0.15 (-0.39, 0.09) | 0.2146 | -0.15 (-0.49, 0.19) | 0.3882 | -0.20 (-0.64, 0.24) | 0.3756 |
| Prescription opioids ($\beta_8$) | | | 0.00 (0.00, 0.00) | 0.5559 | 0.00 (0.00, 0.00) | 0.1725 | 0.00 (0.00, 0.00) | 0.4325 |
| OAT ($\beta_9$) | | | -0.08 (-0.18, 0.02) | 0.1249 | **-0.20 (-0.35, -0.05)** | **0.0103** | **-0.19 (-0.35, -0.03)** | **0.0249** |
| Naloxone kits ($\beta_{10}$) | | | 0.00 (-0.01, 0.02) | 0.7036 | 0.01 (-0.01, 0.02) | 0.2960 | **0.02 (0.01, 0.03)** | **0.0044** |
| **Hospitalizations** |  |  |  |  |  |  |  |  |
| Pre-intervention trend, control ($\beta_1$) | 0.01 (-0.02, 0.04) | 0.5827 | 0.01 (-0.01, 0.03) | 0.4812 | 0.00 (-0.01, 0.02) | 0.6786 | 0.02 (-0.01, 0.04) | 0.1771 |
| Pre-intervention level difference, treated vs. control ($\beta_2$) | 0.58 (-1.28, 2.43) | 0.5438 | 0.58 (-0.87, 2.03) | 0.4391 | 0.30 (-0.30, 0.91) | 0.3305 | 0.26 (-0.24, 0.76) | 0.3071 |
| Pre-intervention trend difference, treated vs. control ($\beta_3$) | 0.01 (-0.01, 0.02) | 0.2655 | 0.01 (0.00, 0.02) | 0.1830 | 0.01 (-0.01, 0.02) | 0.3910 | 0.00 (-0.01, 0.02) | 0.5340 |
| Post-intervention level difference, control ($\beta_4$) | -0.05 (-0.22, 0.11) | 0.5264 | -0.05 (-0.19, 0.09) | 0.4597 | 0.00 (-0.18, 0.17) | 0.9651 | -0.01 (-0.20, 0.18) | 0.9407 |
| Post-intervention trend difference, control ($\beta_5$) | -0.01 (-0.03, 0.02) | 0.5668 | -0.01 (-0.02, 0.01) | 0.3550 | 0.00 (-0.02, 0.01) | 0.7259 | -0.02 (-0.05, 0.02) | 0.3331 |
| Booth-hours ($\beta_6$) | 0.00 (0.00, 0.00) | 0.7313 | 0.00 (0.00, 0.00) | 0.5864 | 0.00 (0.00, 0.00) | 0.9710 | 0.00 (0.00, 0.00) | 0.9016 |
| Post-intervention trend difference, treated vs. control ($\beta_7$) | -0.03 (-0.07, 0.01) | 0.1250 | **-0.03 (-0.05, -0.01)** | **0.0171** | -0.02 (-0.05, 0.01) | 0.1618 | -0.03 (-0.07, 0.01) | 0.2000 |
| Prescription opioids ($\beta_8$) | | | 0.00 (0.00, 0.00) | 0.7570 | 0.00 (0.00, 0.00) | 0.9780 | **0.00 (0.00, 0.00)** | **0.0312** |
| OAT ($\beta_9$) | | | 0.00 (-0.02, 0.01) | 0.6692 | -0.01 (-0.02, 0.01) | 0.5817 | -0.01 (-0.03, 0.01) | 0.3725 |
| Naloxone kits ($\beta_{10}$) | | | 0.00 (0.00, 0.00) | 0.8669 | 0.00 (0.00, 0.00) | 0.8243 | 0.00 (0.00, 0.00) | 0.6703 |
| Deaths |  |  |  |  |  |  |  |  |
| Pre-intervention trend, control ($\beta_1$) | **0.06 (0.04, 0.09)** | **0.0000** | **0.06 (0.05, 0.08)** | **0.0000** | **0.06 (0.04, 0.07)** | **<0.0001** | **0.06 (0.03, 0.08)** | **<0.0001** |
| Pre-intervention level difference, treated vs. control ($\beta_2$) | **1.79 (0.92, 2.66)** | **0.0001** | **1.79 (1.22, 2.36)** | **0.0000** | **0.73 (0.53, 0.93)** | **<0.0001** | **0.37 (0.22, 0.53)** | **<0.0001** |
| Pre-intervention trend difference, treated vs. control ($\beta_3$) | **0.01 (0.00, 0.02)** | **0.0080** | **0.01 (0.01, 0.02)** | **0.0000** | 0.00 (0.00, 0.01) | 0.2468 | **0.00 (-0.01, 0.01)** | **0.8106** |
| Post-intervention level difference, control ($\beta_4$) | -0.01 (-0.15, 0.13) | 0.8591 | -0.01 (-0.08, 0.05) | 0.6998 | 0.01 (-0.12, 0.14) | 0.9325 | -0.02 (-0.18, 0.14) | 0.8366 |
| Post-intervention trend difference, control ($\beta_5$) | **-0.03 (-0.05, -0.01)** | **0.0080** | **-0.03 (-0.04, -0.02)** | **0.0001** | **-0.04 (-0.05, -0.02)** | **<0.0001** | **-0.04 (-0.06, -0.02)** | **0.0022** |
| Booth-hours ($\beta_6$) | 0.00 (0.00, 0.00) | 0.3414 | 0.00 (0.00, 0.00) | 0.2023 | 0.00 (0.00, 0.00) | 0.2466 | 0.00 (0.00, 0.00) | 0.2371 |
| Post-intervention trend difference, treated vs. control ($\beta_7$) | 0.00 (-0.03, 0.04) | 0.8373 | 0.00 (-0.03, 0.03) | 0.8321 | -0.02 (-0.05, 0.02) | 0.3559 | -0.02 (-0.05, 0.02) | 0.3937 |
| Prescription opioids ($\beta_8$) | | | **0.00 (0.00, 0.00)** | **0.0282** | **0.00 (0.00, 0.00)** | **0.0157** | **0.00 (0.00, 0.00)** | **0.0293** |
| OAT ($\beta_9$) | | | **-0.03 (-0.05, -0.02)** | **<0.0001** | **-0.04 (-0.05, -0.03)** | **<0.0001** | **-0.03 (-0.05, -0.02)** | **<0.0001** |
| Naloxone kits ($\beta_{10}$) | | | **0.00 (-0.01, 0.00)** | **0.0005** | 0.00 (0.00, 0.00) | 0.2166 | 0.00 (0.00, 0.00) | 0.8176 |

Restricted models excluded: ED (London, Niagara, Thunder Bay); Hospitalizations (London, Kingston, Niagara, Thunder Bay); Deaths (Niagara, Thunder Bay) based on pre-intervention outcome rates and reported root mean square prediction errors.

The results from the aggregate analysis using the entire observation period (March 2021, inclusive) and restricting to treated PHUs with well-fitting synthetic controls found no effect of booth-hours on ED visits (0.00, 95% CI: -0.01, 0.01; p-value=0.6684), hospitalizations (0.00, 95% CI: 0.00, 0.00; p-value=0.9710) or deaths (0.00, 95% CI: 0.00, 0.00; p-value=0.2466) after controlling for other co-occurring interventions and underlying epidemiological trends. The results were the same when we restricted the observation period to before COVID-19 public health measures were implemented (February 2020, inclusive): 0.00 ED visits (95% CI: -0.01, 0.01; p-value=0.7591), 0.00 hospitalizations (95% CI: 0.00, 0.00; p-value=0.9016), and 0.00 deaths (95% CI: 0.00, 0.00; p-value=0.2371).

Notably, aggregate results revealed a change in epidemiological trend in the synthetic controls (β_5_) for -0.23 ED visits per month (95% CI: -0.35, -0.12; p-value=0.0002) and -0.04 deaths per month (95% CI: -0.05, -0.02; p-value<0.0001); with no additional statistically significant change in trend for treated units (β_7_) for ED visits (−0.15, 95% CI: -0.49, 0.19; p-value=0.3882), hospitalizations (−0.02, 95% CI: -0.05, 0.01; p-value=0.1618) or deaths (−0.02, 95% CI: -0.05, 0.02; p-value=0.3559), respectively. In other words, the aggregate analysis found the same small decline in trend post-implementation across both treated and control units. Finally, aggregate analyses also revealed that OAT reduced ED visits (−0.20, 95% CI: -0.35, -0.05; p-value=0.0103) and deaths (−0.04, 95% CI: -0.05, -0.03; p-value=<0.0001); while prescription opioids for pain management modestly increased deaths (0.0008, 95% CI: 0.0002, 0.0015; p-value=0.0157). These results persisted in our sensitivity analyses.

**Figure 1.**
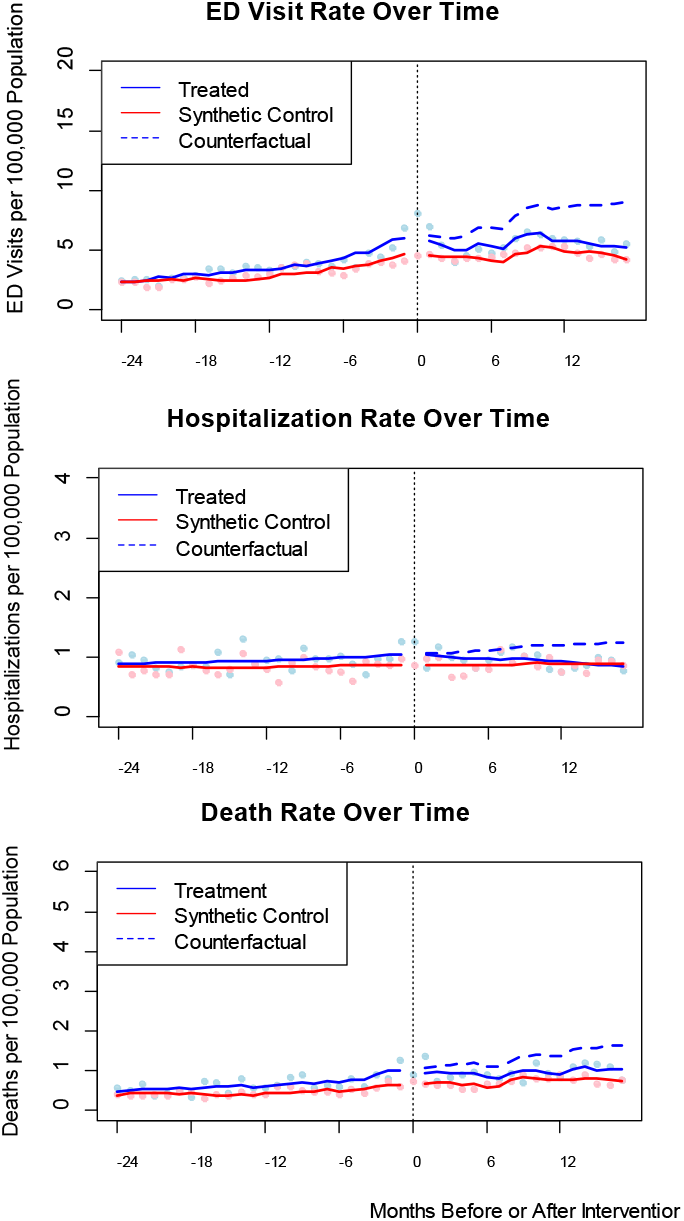
Controlled interrupted time series plots of aggregate analyses restricted to treated PHUs with well-fitted synthetic controls

Results for individual treated/synthetic control PHUs were mixed. Note that many coefficients were assessed relative to the p=0.05 level. There were 102 coefficients (4 coefficients for 3 outcomes across 9 PHUs) that would be described in the following text if their p-value had been below 0.05:

### ED visits

Per booth-hour, ED visits increased in Guelph (0.02, 95% CI: 0.01, 0.03; p-value=0.0059) but declined in Ottawa (−0.0023, 95% CI: -0.0037, -0.0008; p-value=0.0021) and London (−0.02, 95% CI: -0.03, 0.00; p-value=0.0177). Meanwhile, prescription opioids for pain management increased ED visits in London (0.01, 95% CI: 0.00, 0.01; p-value=0.0266), Kingston (0.01, 95% CI: 0.01, 0.01; p-value=0.0001), Thunder Bay (0.0044, 95% CI: 0.0003, 0.0084; p-value=0.0380) and Niagara (0.01, 95% CI: 0.00, 0.01; p-value=0.0463).

Our sensitivity analyses found no effects of booth-hours on ED visits but prescription opioids for pain management decreased ED visits in Toronto (−0.0027, 95% CI: -0.0052, -0.0001; p-value=0.04555), and Ottawa (−0.0039, 95% CI: -0.0059, -0.0019; p-value=0.0003).

### Hospitalizations

There was no statistically significant effect of booth-hours on hospitalizations. However, prescription opioids for pain management increased hospitalizations in Kingston (0.0017, 95% CI: 0.0001, 0.0032; p-value=0.0350) but decreased hospitalizations in Waterloo (−0.0022, 95% CI: -0.0032, -0.0011; p-value=0.0002). Results from the sensitivity analyses found no effect of opioids for pain management on hospitalizations in the period before COVID-19 public health measures were implemented.

### Deaths

Per booth-hour, deaths declined in London (−0.0039, 95% CI: -0.0062, -0.0015; p-value=0.0020), and Thunder Bay (−0.0038, 95% CI: -0.0074, -0.0002; p-value=0.0396). Prescription opioids for pain management increased deaths in London (0.0010, 95% CI: 0.0001, 0.0019; p-value=0.0349). Paradoxically, OAT reduced deaths in London (−0.02, 95% CI: -0.03, -0.01; p-value<0.0001) but increased deaths in Hamilton (0.01, 95% CI: 0.00, 0.03; p-value=0.0169). Naloxone kits reduced deaths in Toronto (−0.0043, 95% CI: -0.01, 0.00) and London (−0.0027, 95% CI: -0.0042, -0.0012; p-value=0.0004).

Our sensitivity analyses found the effects of prescription opioids for pain management (0.0013, 95% CI: 0.0003, 0.0022; p-value=0.0113) and OAT (−0.02, 95% CI: -0.03, -0.01; p-value<0.0001) persisted in London; while OAT (0.01, 95% CI: 0.01, 0.02; p-value=0.0005) and naloxone (0.0077, 95% CI: 0.0022, 0.0131; p-value=0.0074) increased deaths in Kingston. DISCUSSION

Our aggregate analyses found no effects of booth-hours on opioid-related ED visits, hospitalizations, or deaths; but OAT reduced ED visits and deaths. Small, but significant reductions in ED visits (Ottawa, London) and deaths (London, Thunder Bay) were observed locally; while there was an increase in ED visits for Guelph. In other words, 5 of the 36 coefficients for the local effects of booth-hours appeared to be significant at the p=0.05 level.

Our models estimated 18.76 (95% CI: -34.61, -7.50) and 16.47 (95% CI: -28.67, -7.17) ED visits averted per month in Ottawa and London, respectively; and an additional 10.34 (95% CI: 5.17, 15.51) ED visits per month for Guelph. Assuming each visit cost the health care system approximately $304 (Canadian Institute for Health Information, 2020), this equates to $5703 and $5007 saved, and $3134 incurred per month, respectively. Further, we found London’s and Thunder Bay’s CTSs averted 3.21 (95% CI: -5.11, -1.24) and 2.90 (95% CI: -5.63, -0.15) deaths per month, respectively. Using a very modest value of life of $1,000,000 (Griffiths & Vadlamudi, 2016) this equates to $3,210,000 and $2,900,000 saved monthly. However, we *do not* encourage limiting cost-effectiveness or cost-benefit exercises to overdose events averted since costs of operating OPSs, for example, while minimal (e.g. $20,367/month each, 40 hours/week) still include other program, administrative, phone, data management and IT expenses that are not specific to these outcomes (Ministry of Health and Long-Term Care, 2018a, 2018b). To better determine if OPS/CTSs are a cost-effective or even cost-saving intervention, we recommend incorporating costs averted from reduced transmission of blood borne infections and other drug use injuries (i.e. skin, soft tissue, and vascular infections and their sequelae) (Panagiotoglou et al., 2021).

Extensive research on the effects of supervised consumption sites (and their variants) has yet to show consistent population-level benefits. Until recently, all the evidence on the effectiveness of supervised consumption sites in Canada was based on Vancouver’s Insite. First implemented in September 2003, Insite is in the Downtown Eastside – a unique neighborhood with one of the highest concentrations of people who use illicit drugs globally (Hayashi et al., 2019). Numerous studies have explored Insite’s effectiveness on drug related harms and acceptability in the community. Among the most rigorous of these, a study led by Marshall et al. demonstrated Insite decreased the local fatal overdose rate by 35% from 253.1 to 165.1 deaths per 100,000 person-years (Marshall BDL, Milloy MJ, Wood E, Montaner JS, & Kerr T, 2011). Differences in magnitude between Marshall et al.’s findings and ours may be a consequence of several important methodological differences. Their analysis compared overdose mortality rates for the population within 500m of Insite to the rest of the city’s population pre-(1 January 2001 to 20 September 2003) and post-implementation (21 September 2005 to 31 December 2005). While innovative at the time, this seminal paper did not account for underlying epidemiological trends or other co-occurring and time-varying interventions, nor restricted comparison to neighborhoods comparable to the Downtown Eastside, potentially overestimating the effect of Insite on fatal overdose rates.

More recently, a mathematical model of the individual and synergistic effects of British Columbia’s harm reduction interventions (OAT, take home naloxone, and OPS) found all three play a significant role in stemming overdose deaths, with OPS averting 290 deaths (credible interval (CrI): 160 – 350) over 21 months (Michael A. Irvine et al., 2019). Differences in the effect sizes between their findings and ours may be, in part, owing to a key assumption underlying their model: in the absence of medical or peer intervention each overdose results in death (M. A. Irvine et al., 2018; Michael A. Irvine et al., 2019). While overdoses do cause life-threatening bradypnea (also referred to as opioid induced respiratory depression, classified as < 12 breaths per minute), respiratory distress, particularly when mild, can spontaneously resolve (Ballantyne, 2007; Toronto Public Health, 2022). In fact, in Irvine et al.’s 2019 paper, although the baseline death rate was set to 1.0 with large credible intervals, posterior estimates showed this to be much smaller (0.0682, 95% CrI: 0.0663 – 0.0701) (Michael A. Irvine et al., 2019). While the authors do not speak to this discrepancy directly in the 2019 paper, they do *briefly* acknowledge a large difference between the prior and posterior estimates in a prior paper examining the effects of BC’s take home naloxone program (M. A. Irvine et al., 2018). This uncertainty around the baseline death rate per overdose event coupled with differences in population substance use preferences, patterns and geographic diffusion; extent of fentanyl contamination of the illicit drug supply; observation period (i.e. the BC paper examined the effects of these interventions when most services *and* overdose events remained concentrated within Vancouver’s Downtown Eastside); and delivery models of OPS/CTSs may further explain why Irvine et al. observed substantially larger effect sizes for OPSs in British Columbia than we do here.

Our study is not the first to find a lack of effect of supervised consumption sites on mortality, but it is the largest. An evaluation of Sydney, Australia’s supervised injection centre (NCHECR, 2007) and another of OPSs in mid-sized communities on Vancouver Island (The Evaluation of Overdose Prevention Sites Working Group & Lori Wagar, 2018) both noted a decrease in local paramedic attendance, but no effect on overdose related mortality. The authors of these reports considered the rarity of fatal events and subsequent challenges with study power as possible explanations for the absence of a measurable effect; and specific to Sydney, a co-occurring decline in the local heroin supply.

Our results also agree with simpler analysis we conducted to evaluate British Columbia’s recently implemented OPSs (Panagiotoglou, 2022). In both studies, we used controlled interrupted time series and reported aggregate, population-level results. However, in the BC study we were not able to control for other time-varying interventions nor the intensity of the intervention itself. Despite these differences, both studies found a lack of effect on hospitalization and mortality rates. The BC analysis found OPSs reduced ED visits whereas our Ontario analysis did not. This may be due to differences in provincial OPS guidelines for medical care follow-up after on-site overdose reversal (BC Centre on Substance Use, 2017). Finally, both studies found a decline in the death rate trend in their respective aggregate control populations which was mirrored in the treated units (Panagiotoglou, 2022). While the cause of this decline in underlying epidemiological trend is not clear, it may be a combination of gradual changes in opioid drug use behaviour (e.g. prioritizing using substances with a peer, testing substances for contaminants), the public’s perception of substance use disorder, and law enforcement activity. The effects of this cultural shift may be imperceptibly small at the local treated/control unit level, but discernable in the aggregate analyses, and worth additional exploration.

This common change in trend for the treated/control unit dyads lends additional credence to the comparability of the groups. Alternatively, a change in the control’s trend that was not also observed in the treated unit would indicate substantial and problematic differences between the populations.

The lack of observed effect of OPS/CTSs on our outcomes may be a consequence of access barriers first described elsewhere. Work examining successful implementation and uptake of OPSs in British Columbia found persisting stigma and police presence reduced their social acceptability and use by at-risk populations (Collins et al., 2019). Meanwhile, hours of operation, facility capacity, and absence of safe inhalation rooms limited their effectiveness (The Evaluation of Overdose Prevention Sites Working Group & Lori Wagar, 2018). Similar barriers have been described in qualitative work on Ontario’s OPSs (Foreman-Mackey, Bayoumi, Miskovic, Kolla, & Strike, 2019). This may also explain why OAT reduces deaths, since patients receiving OAT have already overcome challenges to access.

Despite concerns that harm reduction interventions may encourage risky behaviour (Hedlund, 2000) and therefore explain the lack of observed effects in our study, a team examining the prevalence of unintentional overdose in a longitudinal cohort of people who inject drugs in Toronto has found no evidence of risk compensation (Scheim et al., 2021). Finally, while drug use and fentanyl consumption, in particular, have increased since March 2020 (Statistics Canada, 2021), these consumption changes do not explain the lack of observed effects in our sensitivity analyses. Together, our results suggest the effectiveness of OPS/CTSs is more modest than described elsewhere, and/or context specific.

#### Strengths and limitations

Our study had some limitations. We used reported hours of operation gleaned from OPS/CTS websites, online platforms (i.e. Facebook and Twitter) or reported by local media to estimate booth-hours provided per month. Where information was missing, we assumed the number of booths matched the number included in public reports or plans prior to opening. Despite exhaustive searches, we were unable to confirm the number of booth-hours provided at Toronto’s The Works’ two recently implemented hotel-based resident sites, and for Ottawa’s mobile site, and did not include these sites in our monthly booth-hour estimates. For COVID-19 related changes to services, if no explicit update on the number of booths was available, we assumed the number was adjusted to meet public health guidelines (e.g. where three booths normally operated side-by-side, we assumed the middle booth was temporarily unavailable until guidelines were revised) (Public Health Ontario, 2020). These assumptions and exclusions may underestimate the number of booth-hours per PHU and overestimate the marginal effect of each additional booth-hour. However, they do not detract from the overall observed effect that booth-hours had no effect on ED visit, hospitalization or death rates in our aggregate analyses.

With respect to naloxone kits distributed by community-based organizations (separate from those dispensed by participating pharmacies), we used annual reports to estimate monthly counts whenever more granular data were not available (e.g. distribution campaign or blitz for a specific period). Again, this likely introduced error to the month-to-month number of kits distributed but should have minimal effect on the overall impact of naloxone kits readily available for private use.

While we cannot definitively rule out that travel from control to treated PHUs did not dilute treatment effects, using maps to confirm OPS/CTSs were well within PHU boundaries and assuming distance-decay effects described in other studies apply here (Marshall BDL et al., 2011; The Evaluation of Overdose Prevention Sites Working Group & Lori Wagar, 2018), it is unlikely that population mobility across boundaries explains the absence of observed effects. This lack of evidence of spillover effects satisfies a critical assumption of synthetic control methods (Bouttell et al., 2018).

The response to the COVID-19 pandemic introduced some challenges for our study, including delays in public reporting of outcome data by Ontario Public Health and public health units (e.g. naloxone kits distributed). However, we were still able to observe at least fourteen months (and over three years for early adopters Toronto and Ottawa) of intervention effect post-implementation after allowing for a three month intervention ramp up period as recommended by others (The Evaluation of Overdose Prevention Sites Working Group & Lori Wagar, 2018). Further, by including periods after federal/provincial COVID-19 emergency orders were implemented in March 2020, we were able to test the effects of booth-hours with dramatic changes in capacity within distinct populations (Box, Jenkins, & Reinsel, 2011). Although we were able to adjust for reduced service capacity and changes in naloxone distribution, and OAT and prescription opioids for pain management dispensing because of social distancing measures, we were unable to directly adjust for changes in toxicity of the illicit drug supply following border closures. Potential increases in drug toxicity along with temporary reductions in booth-hours and clients’ hesitation to use services may explain the spikes in ED visit and mortality rates observed immediately after March 2020 (Supplementary Figures 1 and 3, respectively) (Beard et al., 2019; Statistics Canada, 2021).

Finally, we were not able to create sufficiently well-fitted synthetic controls for all our treated PHUs across all three outcomes. Some of this was anticipated given that outcome rates for Niagara and Thunder Bay were outside the convex hull of rates observed in the donor pool in the six months leading to the implementation of the intervention. To work around this, we reported the individual effects between treated and synthetic controls for all pairs in the supplement (Table 3). For our main results, we reported the findings from our aggregate analyses restricted to treated PHU/synthetic control pairs where there was no observed pre-intervention trend differences between the two and whose RMSPE was small (n_ED_=6, n_Hosp_=5, and n_Deaths_=7). We also reported for the reader the aggregate results including all treated PHU/synthetic control pairs (n=9). While likely biased, these results also found no effect of booth-hours on our outcomes. Although novel approaches to address the challenges of pre-treatment fit such as Augmented Synthetic Control Methods are becoming available, we opted to combine the traditional synthetic controls as first developed by Abadie et al. with interrupted time series methods to maximize the accessibility of this study (Ben-Michael, Feller, & Rothstein, 2021).

By comparing outcome rates between treated PHUs and synthetic controls with excellent fit, controlling for concomitant time-varying interventions, conducting extensive sensitivity analyses, pooling the data using a multiple baseline approach for aggregate analyses, and comparing the plausibility of our results with site specific use statistics – ours is the first rigorous, province-wide study on the effects of supervised consumption sites on a population outside British Columbia. Our results suggest that the effects of OPS/CTSs are modest and/or context specific at best.

## CONCLUSION

Supervised consumption sites are among a set of harm reduction interventions increasingly implemented across Canada to stem the ongoing opioid overdose epidemic. Our study found no effect of booth-hours on ED visit, hospitalization or deaths across Ontario, and small local effects. Although OPS/CTSs do not appear in this study to have beneficial population level effects on overdose-related ED visits, hospitalizations or deaths, they may improve access to other critical services not captured here (e.g. primary care, mental health, housing/social services) and reduce infections and injuries from illicit drug use.

## Data Availability

All data produced in the present study are available upon reasonable request to the authors. Data used are also available across multiple public sources including Ontario Public Health's Opioid Tool, the Ontario Drug Policy and Research Network and public health units' annual reports or dashboards.

## Supplementary Material

**Table 1.**
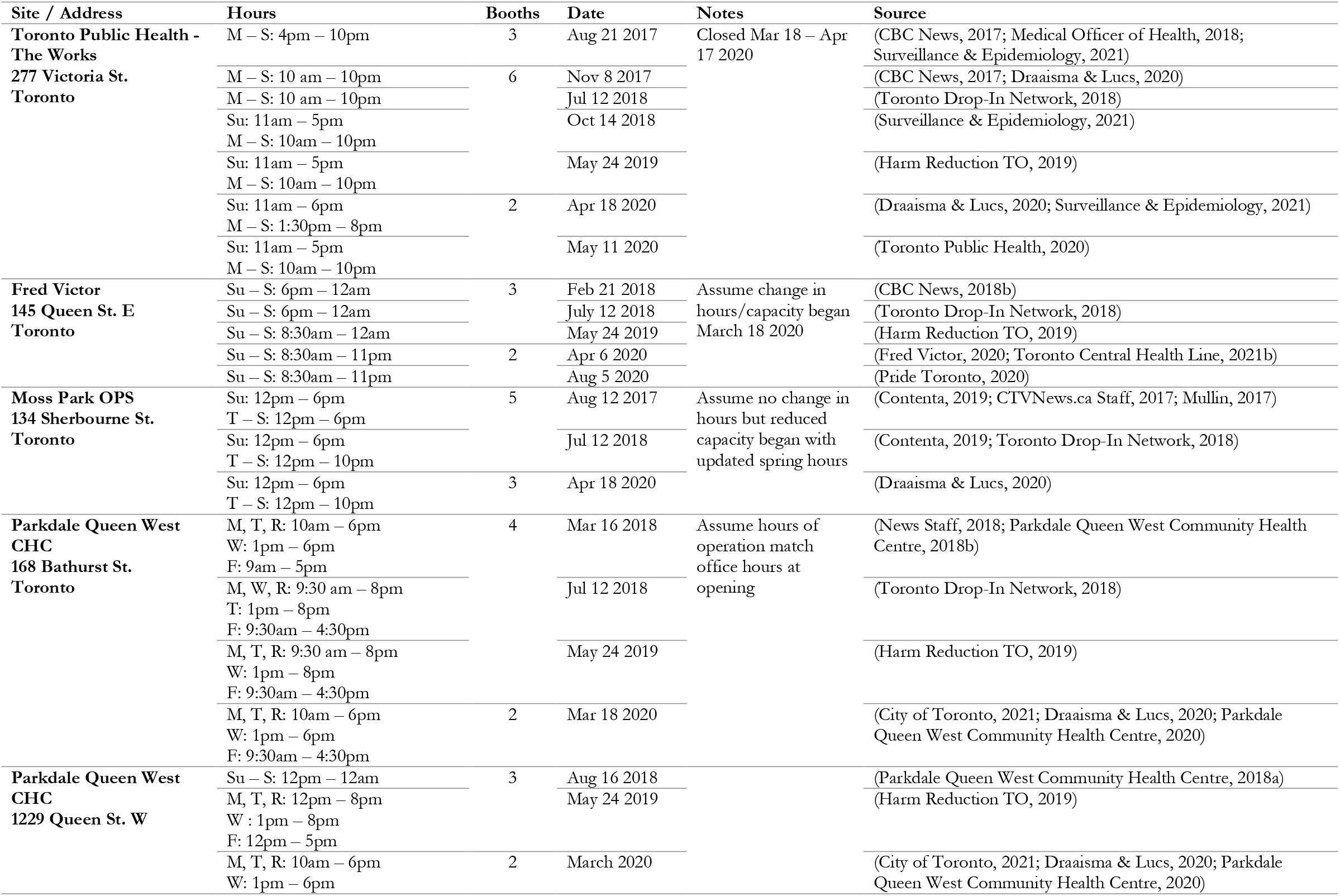
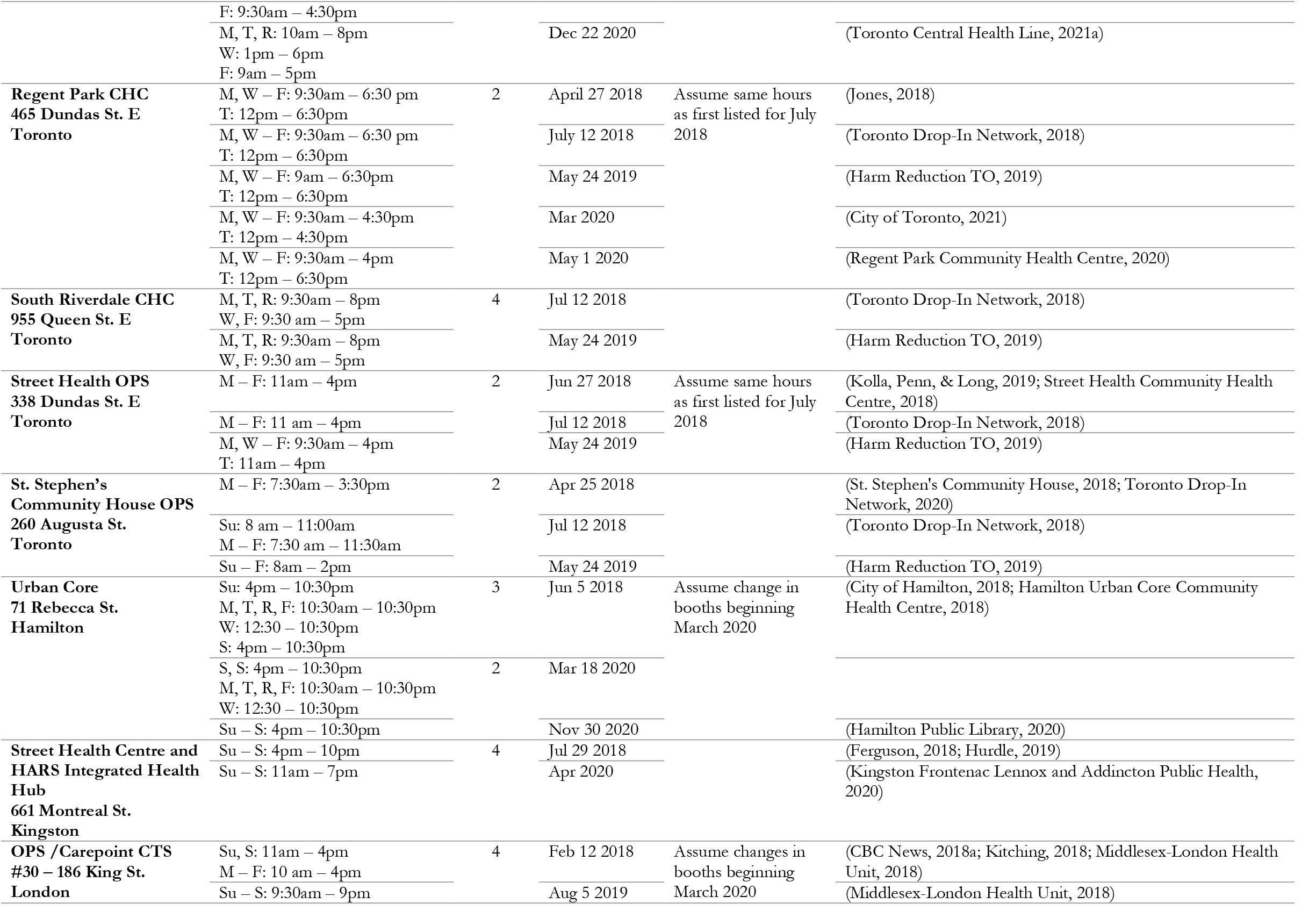
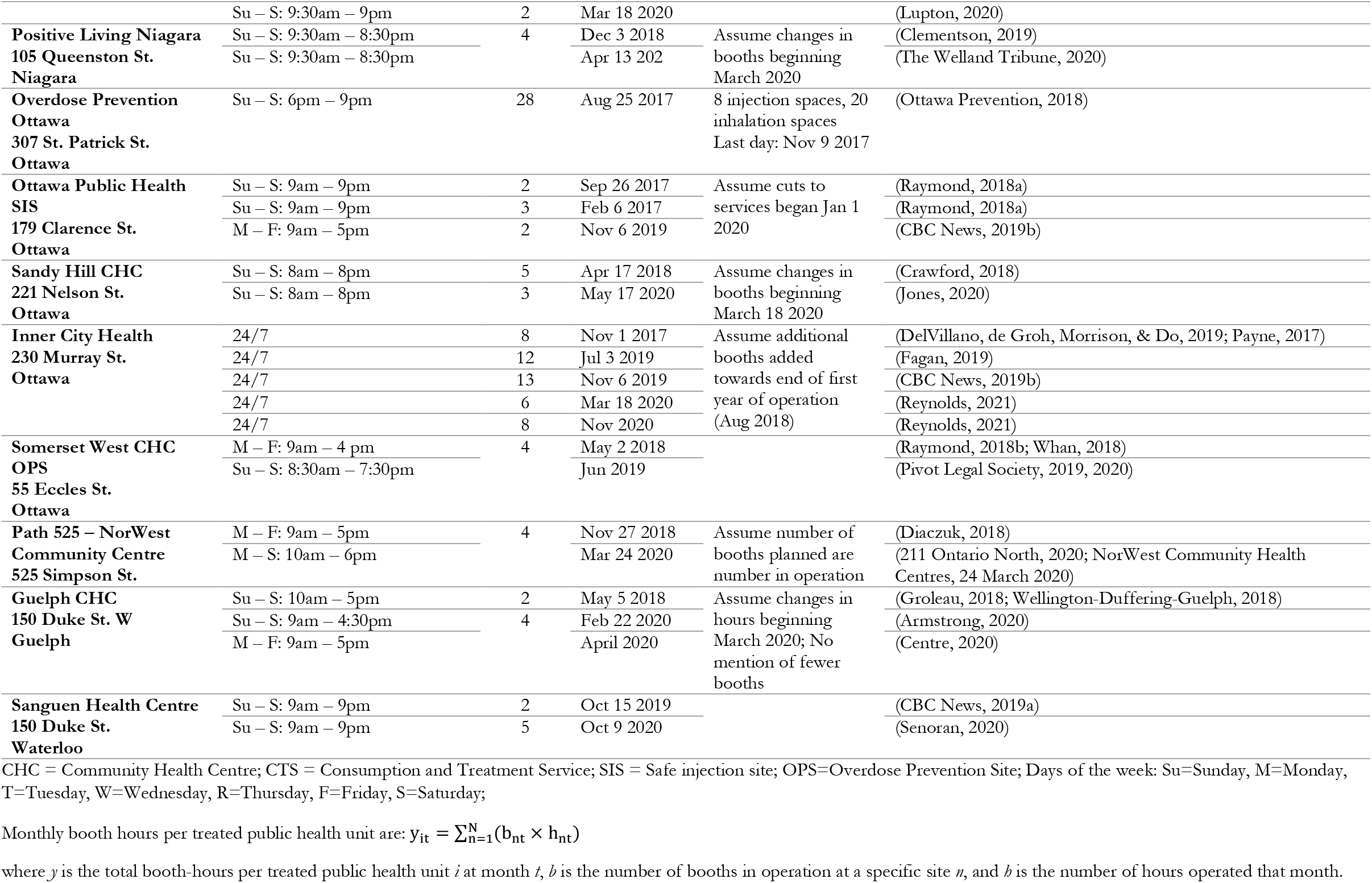
Overdose Prevention Site and Consumption and Treatment Services hours of operation and booths/spaces available

**Table 2.**
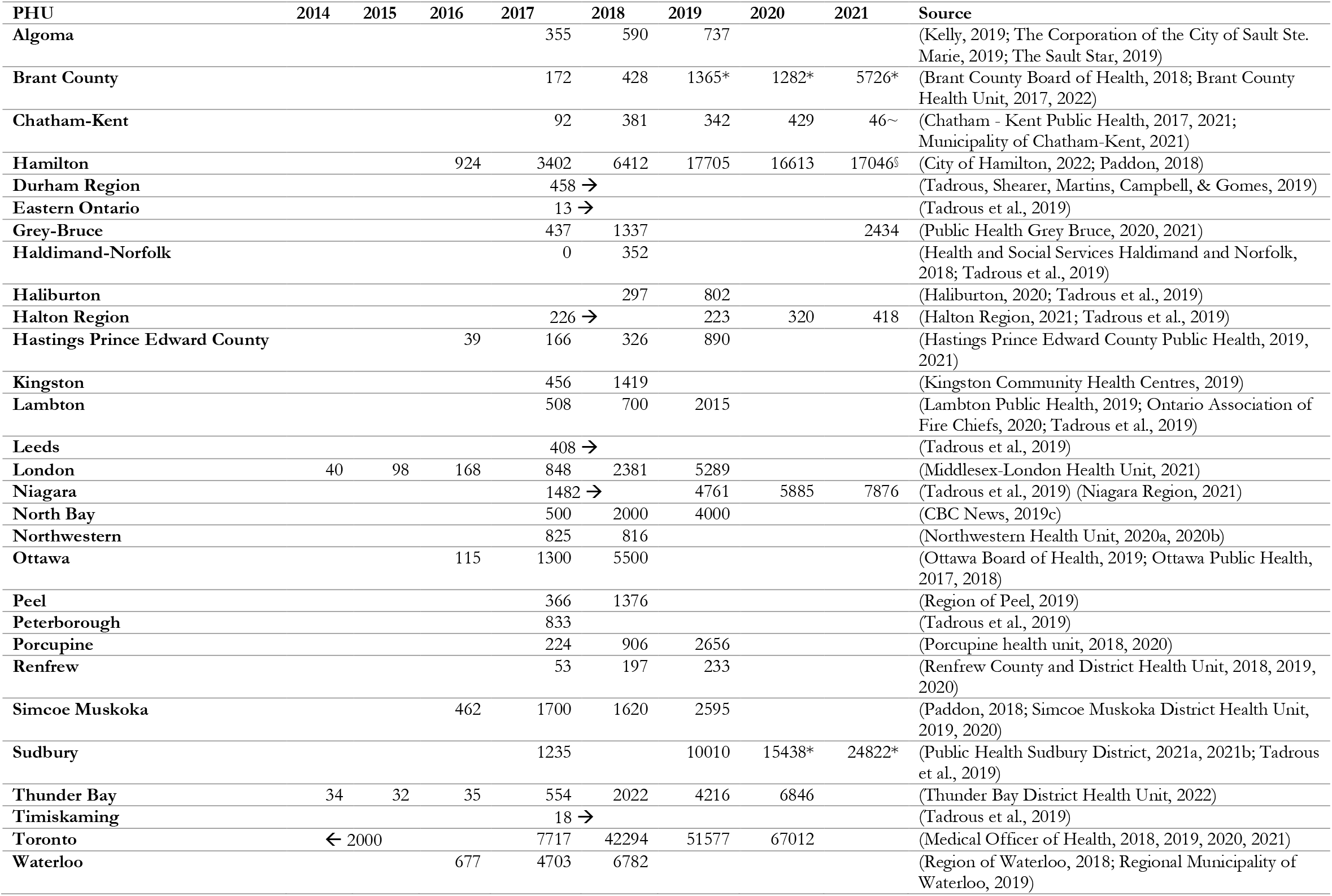
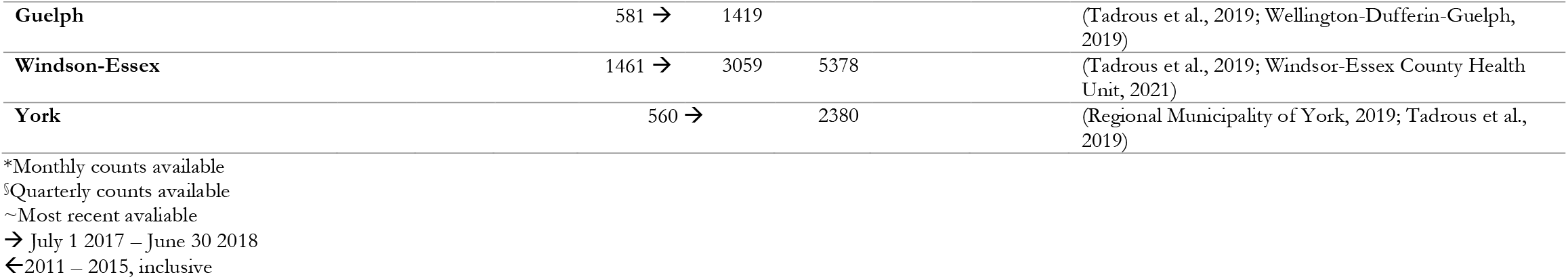
Naloxone kits dispensed by Public Health Units and community-based organizations

**Table 3.**
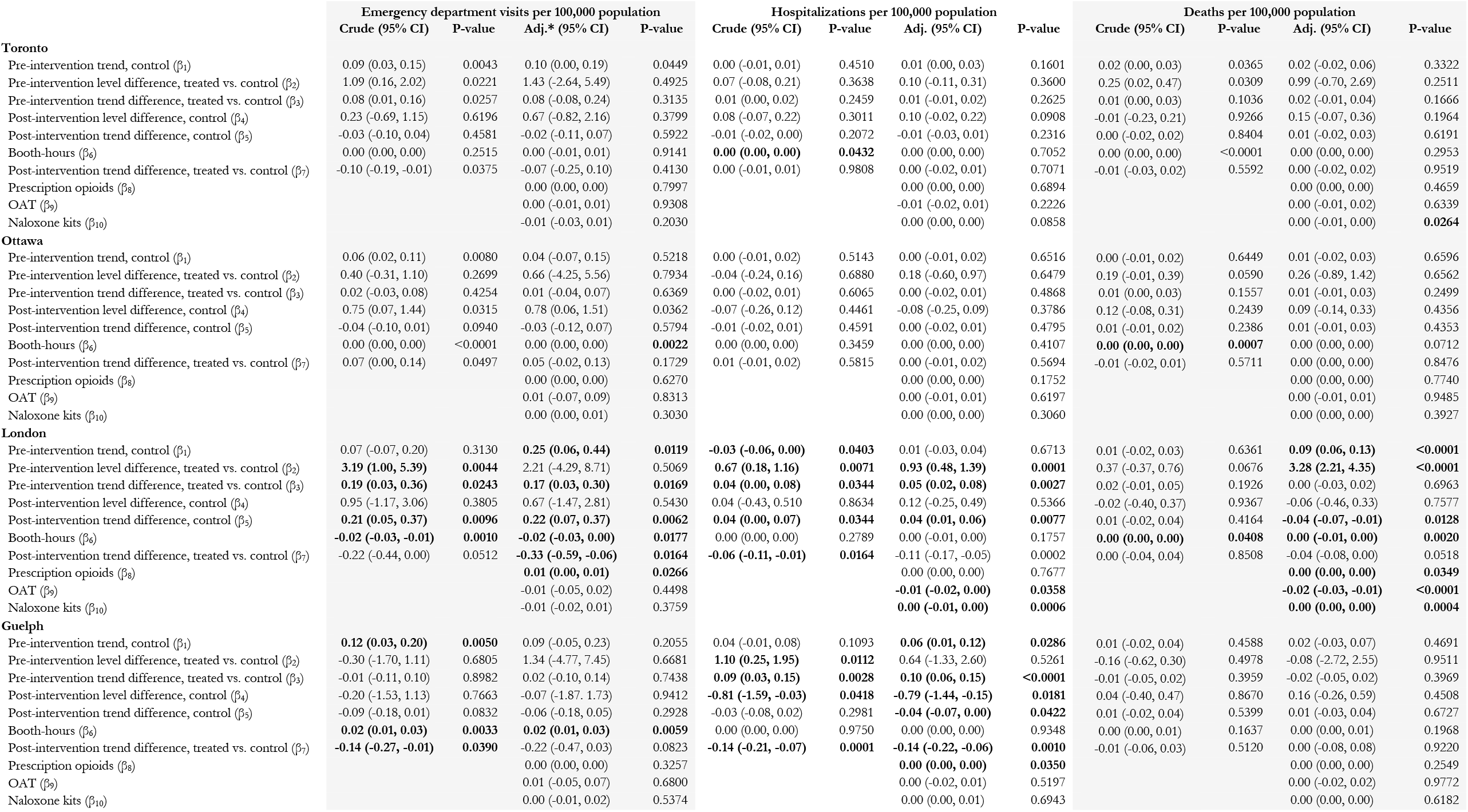
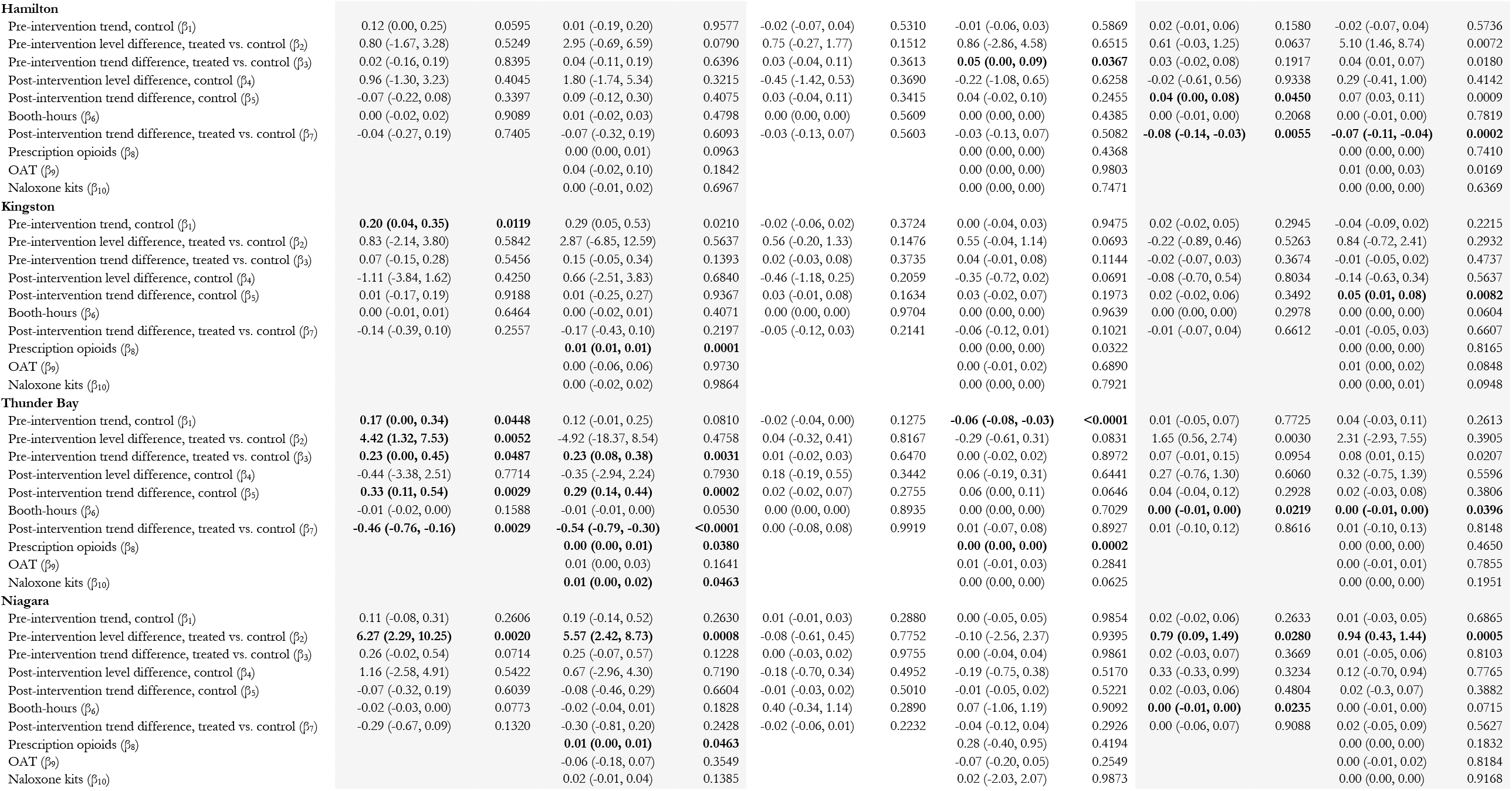
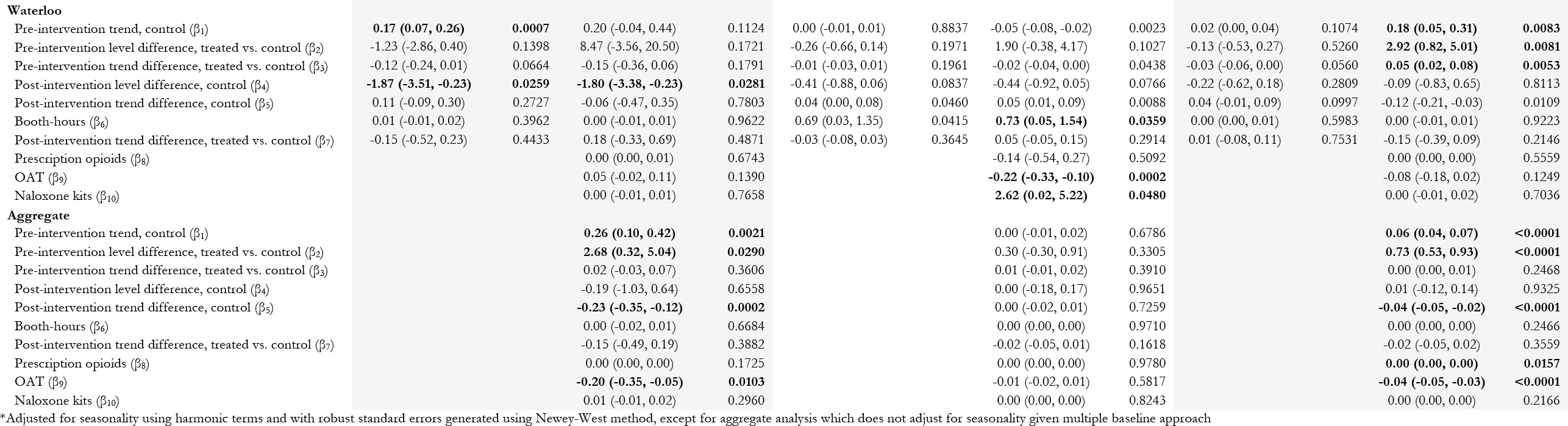
Crude and adjusted segmented regression output for treated/synthetic control unit pairs and aggregate analysis

**Figure 1.**
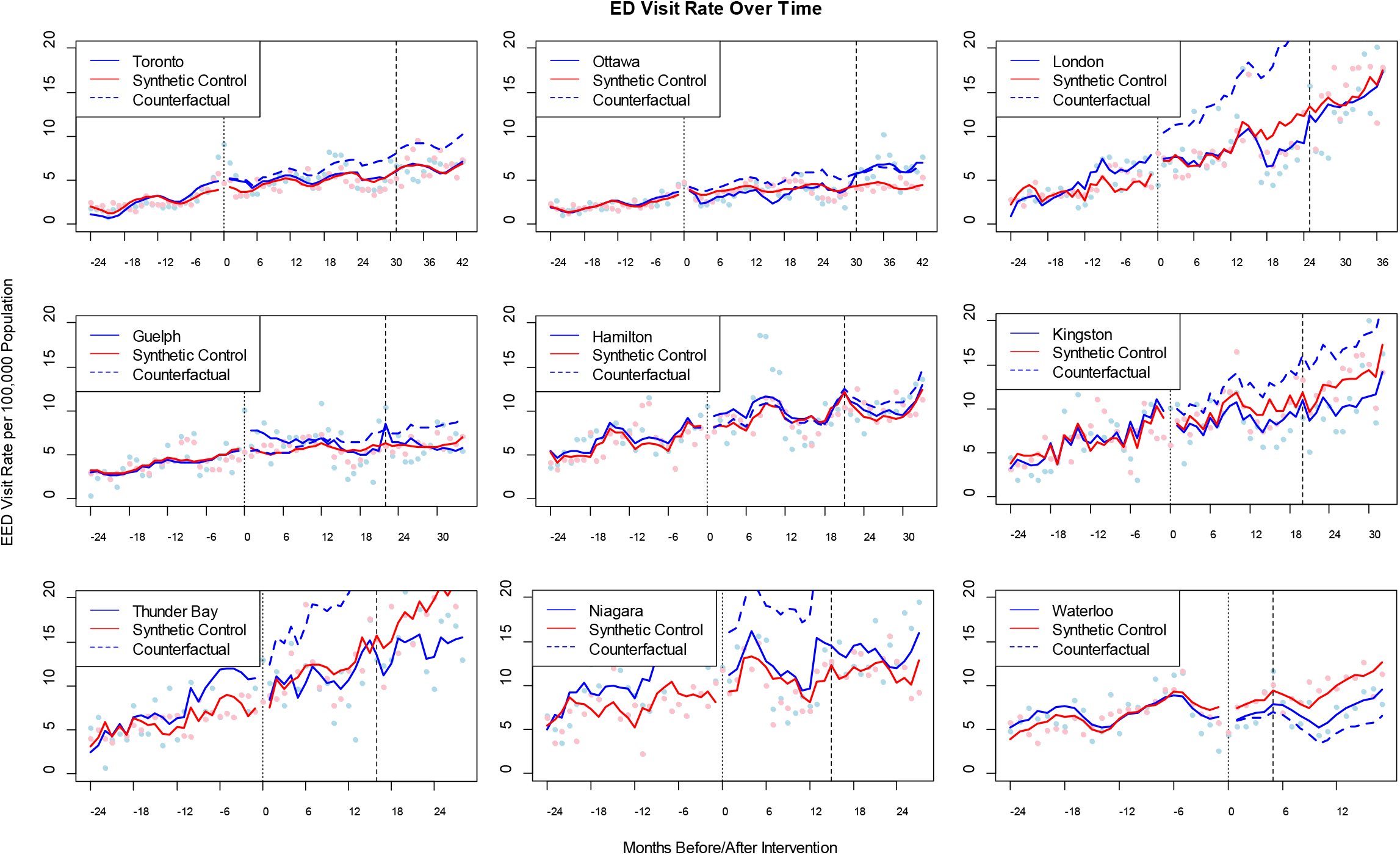
Controlled interrupted time-series of emergency department (ED) visits plotted per treated public health unit – synthetic control pair, with counterfactuals; Dark vertical dashed line = March 2020; Counterfactual represents the expected outcome rates where the intervention set to 0 in the regression models.

**Figure 2.**
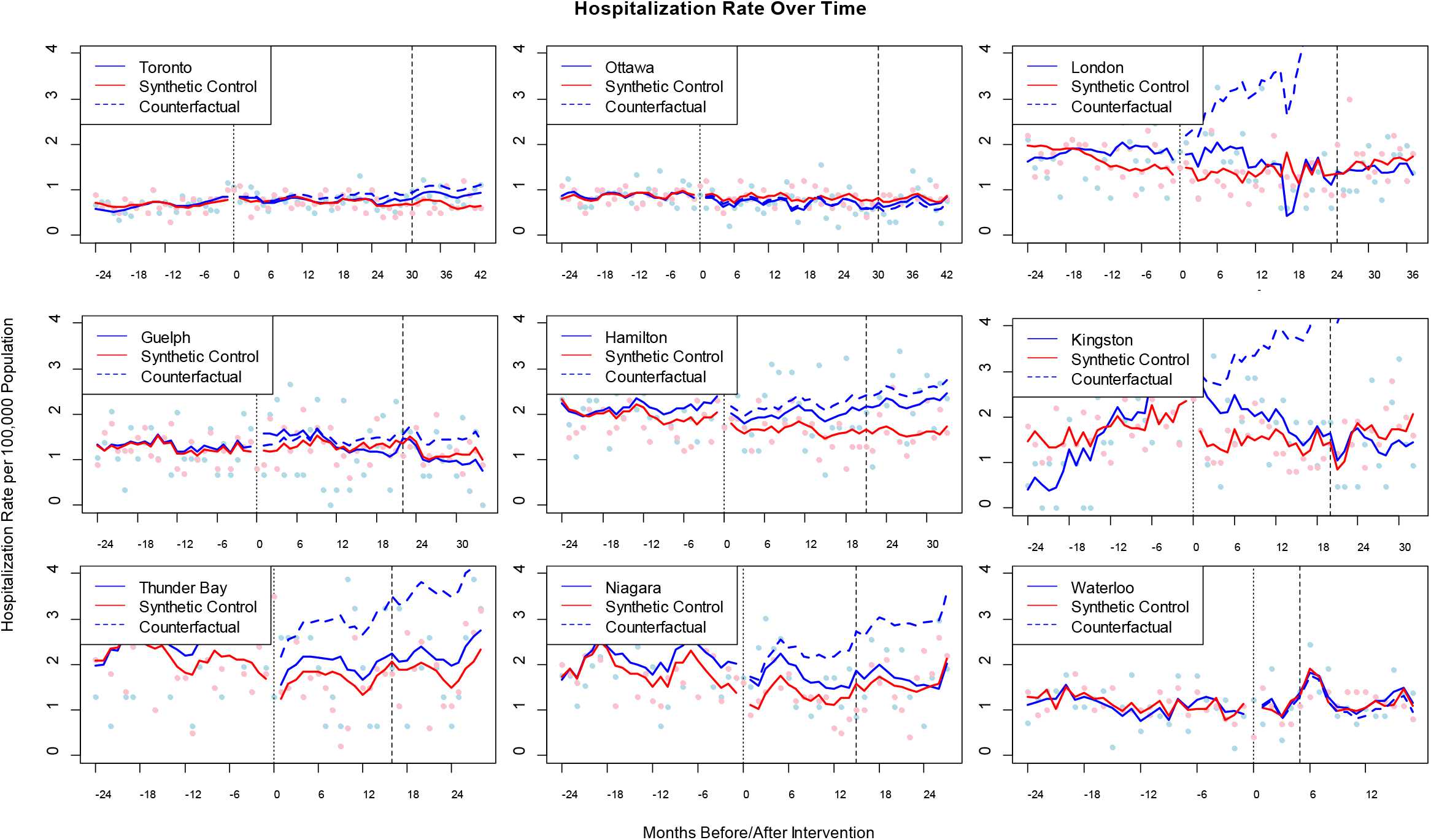
Controlled interrupted time-series of hospitalizations plotted per treated public health unit – synthetic control pair, with counterfactuals; Dark vertical dashed line = March 2020; Counterfactual represents the expected outcome rates were the intervention set to 0 in the regression models.

**Figure 3.**
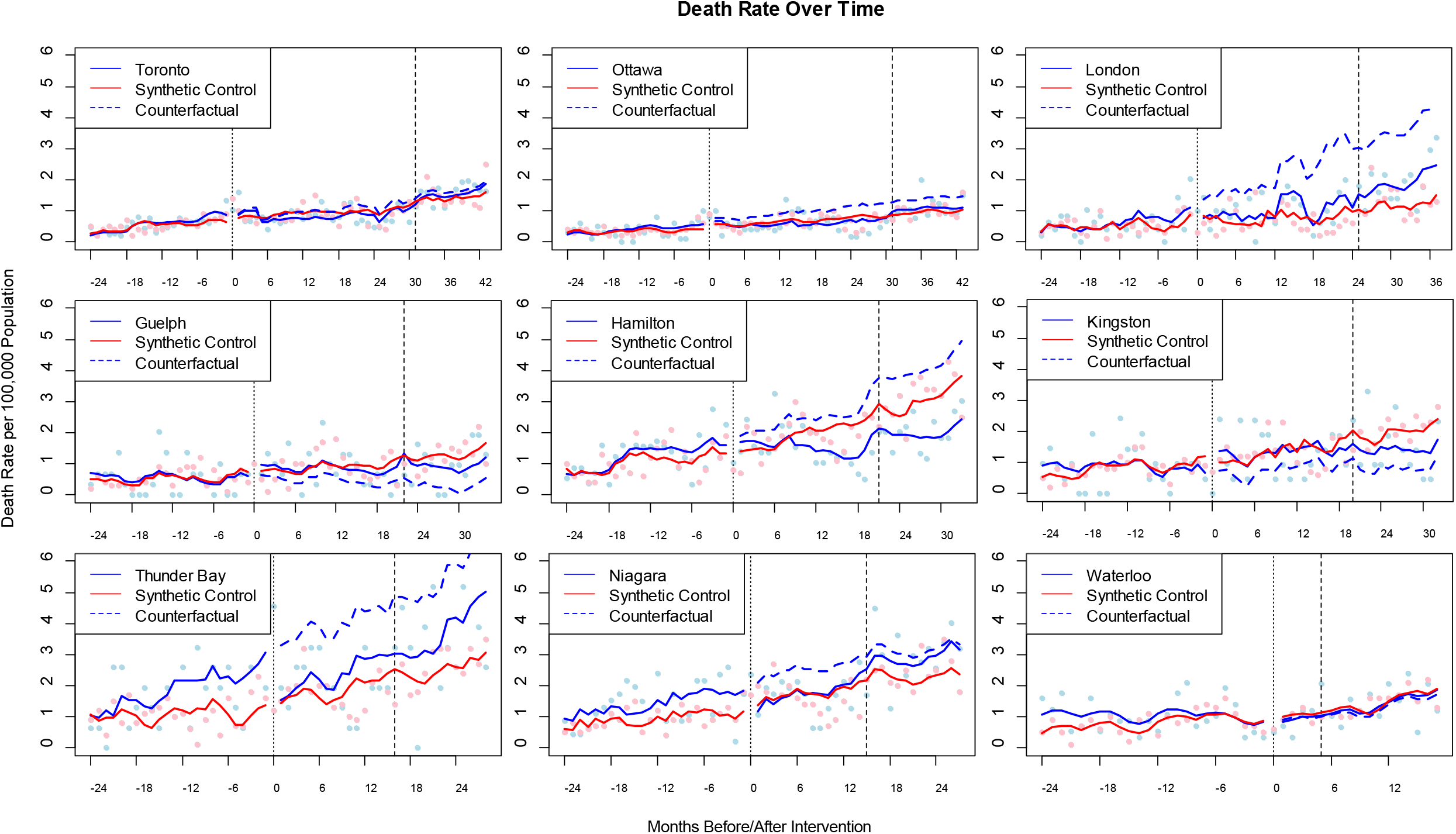
Controlled interrupted time-series of mortalities plotted per treated public health unit – synthetic control pair, with counterfactuals; Dark vertical dashed line = March 2020; Counterfactual represents the expected outcome rates where the intervention set to 0 in the regression models.

