## Supplementary tables and figures for "Using synthetic controls to estimate the population-level effects of Ontario’s recently implemented overdose prevention sites and consumption and treatment services" (from another version of this preprint: version 2, 2022-04-29)

**Table of Contents**

Table 1. Overdose Prevention Site and Consumption and Treatment Services hours of operation and booths/spaces available (pg. 2)

Table 2. Naloxone kits dispensed by Public Health Units and community-based organizations (pg. 5)

Table 3. Crude and adjusted segmented regression output for treated/synthetic control unit pairs and aggregate analysis (pg. 6)

Figure 1. Controlled interrupted time-series of opioid-related ED visits plotted per treated public health unit – synthetic control pair, with counterfactual (pg. 9)

Figure 2. Controlled interrupted time-series of opioid-related hospitalizations plotted per treated public health unit – synthetic control pair, with counterfactual (pg. 10)

Figure 3. Controlled interrupted time-series of opioid-related mortalities plotted per treated public health unit – synthetic control pair, with counterfactual (pg. 11)

Supplement Data Sources (pg. 12)

Table 1. Overdose Prevention Site and Consumption and Treatment Services hours of operation and booths/spaces available

| Site / Address | Hours | Booths | Date | Notes | Source |
| --- | --- | --- | --- | --- | --- |
| Toronto Public Health - The Works  277 Victoria St.  Toronto | M – S: 4pm – 10pm | 3 | Aug 21 2017 | Closed Mar 18 – Apr 17 2020 | [1-3] |
|  | M – S: 10 am – 10pm | 6 | Nov 8 2017 |  | [2, 4] |
|  | M – S: 10 am – 10pm |  | Jul 12 2018 |  | [5] |
|  | Su: 11am – 5pm  M – S: 10am – 10pm |  | Oct 14 2018 |  | [3] |
|  | Su: 11am – 5pm  M – S: 10am – 10pm |  | May 24 2019 |  | [6] |
|  | Su: 11am – 6pm  M – S: 1:30pm – 8pm | 2 | Apr 18 2020 |  | [3, 4] |
|  | Su: 11am – 5pm  M – S: 10am – 10pm |  | May 11 2020 |  | [7] |
| Fred Victor  145 Queen St. E  Toronto | Su – S: 6pm – 12am | 3 | Feb 21 2018 | Assume change in hours/capacity began March 18 2020 | [8] |
|  | Su – S: 6pm – 12am |  | July 12 2018 |  | [5] |
|  | Su – S: 8:30am – 12am |  | May 24 2019 |  | [6] |
|  | Su – S: 8:30am – 11pm | 2 | Apr 6 2020 |  | [9, 10] |
|  | Su – S: 8:30am – 11pm |  | Aug 5 2020 |  | [11] |
| Moss Park OPS  134 Sherbourne St.  Toronto | Su: 12pm – 6pm  T – S: 12pm – 6pm | 5 | Aug 12 2017 | Assume no change in hours but reduced capacity began with updated spring hours | [12-14] |
|  | Su: 12pm – 6pm  T – S: 12pm – 10pm |  | Jul 12 2018 |  | [5, 13] |
|  | Su: 12pm – 6pm  T – S: 12pm – 10pm | 3 | Apr 18 2020 |  | [4] |
| Parkdale Queen West CHC  168 Bathurst St.  Toronto | M, T, R: 10am – 6pm  W: 1pm – 6pm  F: 9am – 5pm | 4 | Mar 16 2018 | Assume hours of operation match office hours at opening | [15, 16] |
|  | M, W, R: 9:30 am – 8pm  T: 1pm – 8pm  F: 9:30am – 4:30pm |  | Jul 12 2018 |  | [5] |
|  | M, T, R: 9:30 am – 8pm  W: 1pm – 8pm  F: 9:30am – 4:30pm |  | May 24 2019 |  | [6] |
|  | M, T, R: 10am – 6pm  W: 1pm – 6pm  F: 9:30am – 4:30pm | 2 | Mar 18 2020 |  | [4, 17, 18] |
| Parkdale Queen West CHC  1229 Queen St. W  Toronto | Su – S: 12pm – 12am | 3 | Aug 16 2018 |  | [19] |
|  | M, T, R: 12pm – 8pm  W : 1pm – 8pm  F: 12pm – 5pm |  | May 24 2019 |  | [6] |
|  | M, T, R: 10am – 6pm  W: 1pm – 6pm  F: 9:30am – 4:30pm | 2 | March 2020 |  | [4, 17, 18] |
|  | M, T, R: 10am – 8pm  W: 1pm – 6pm  F: 9am – 5pm |  | Dec 22 2020 |  | [20] |
| Regent Park CHC  465 Dundas St. E  Toronto | M, W – F: 9:30am – 6:30 pm  T: 12pm – 6:30pm | 2 | April 27 2018 | Assume same hours as first listed for July 2018 | [21] |
|  | M, W – F: 9:30am – 6:30 pm  T: 12pm – 6:30pm |  | July 12 2018 |  | [5] |
|  | M, W – F: 9am – 6:30pm  T: 12pm – 6:30pm |  | May 24 2019 |  | [6] |
|  | M, W – F: 9:30am – 4:30pm  T: 12pm – 4:30pm |  | Mar 2020 |  | [17] |
|  | M, W – F: 9:30am – 4pm  T: 12pm – 6:30pm |  | May 1 2020 |  | [22] |
| South Riverdale CHC  955 Queen St. E  Toronto | M, T, R: 9:30am – 8pm  W, F: 9:30 am – 5pm | 4 | Jul 12 2018 |  | [5] |
|  | M, T, R: 9:30am – 8pm  W, F: 9:30 am – 5pm |  | May 24 2019 |  | [6] |
| Street Health OPS  338 Dundas St. E  Toronto | M – F: 11am – 4pm | 2 | Jun 27 2018 | Assume same hours as first listed for July 2018 | [23, 24] |
|  | M – F: 11 am – 4pm |  | Jul 12 2018 |  | [5] |
|  | M, W – F: 9:30am – 4pm  T: 11am – 4pm |  | May 24 2019 |  | [6] |
| St. Stephen’s Community House OPS  260 Augusta St.  Toronto | M – F: 7:30am – 3:30pm | 2 | Apr 25 2018 |  | [25, 26] |
|  | Su: 8 am – 11:00am  M – F: 7:30 am – 11:30am |  | Jul 12 2018 |  | [5] |
|  | Su – F: 8am – 2pm |  | May 24 2019 |  | [6] |
| Urban Core  71 Rebecca St.  Hamilton | Su: 4pm – 10:30pm  M, T, R, F: 10:30am – 10:30pm  W: 12:30 – 10:30pm  S: 4pm – 10:30pm | 3 | Jun 5 2018 | Assume change in booths beginning March 2020 | [27, 28] |
|  | S, S: 4pm – 10:30pm  M, T, R, F: 10:30am – 10:30pm  W: 12:30 – 10:30pm | 2 | Mar 18 2020 |  |  |
|  | Su – S: 4pm – 10:30pm |  | Nov 30 2020 |  | [29] |
| Street Health Centre and HARS Integrated Health Hub  661 Montreal St.  Kingston | Su – S: 4pm – 10pm | 4 | Jul 29 2018 |  | [30, 31] |
|  | Su – S: 11am – 7pm |  | Apr 2020 |  | [32] |
| OPS /Carepoint CTS  #30 – 186 King St.  London | Su, S: 11am – 4pm  M – F: 10 am – 4pm | 4 | Feb 12 2018 | Assume changes in booths beginning March 2020 | [33-35] |
|  | Su – S: 9:30am – 9pm |  | Aug 5 2019 |  | [33] |
|  | Su – S: 9:30am – 9pm | 2 | Mar 18 2020 |  | [36] |
| Positive Living Niagara  105 Queenston St.  Niagara | Su – S: 9:30am – 8:30pm | 4 | Dec 3 2018 | Assume changes in booths beginning March 2020 | [37] |
|  | Su – S: 9:30am – 8:30pm |  | Apr 13 202 |  | [38] |
| Overdose Prevention Ottawa  307 St. Patrick St.  Ottawa | Su – S: 6pm – 9pm | 28 | Aug 25 2017 | 8 injection spaces, 20 inhalation spaces  Last day: Nov 9 2017 | [39] |
| Ottawa Public Health SIS  179 Clarence St.  Ottawa | Su – S: 9am – 9pm | 2 | Sep 26 2017 | Assume cuts to services began Jan 1 2020 | [40] |
|  | Su – S: 9am – 9pm | 3 | Feb 6 2017 |  | [40] |
|  | M – F: 9am – 5pm | 2 | Nov 6 2019 |  | [41] |
| Sandy Hill CHC  221 Nelson St.  Ottawa | Su – S: 8am – 8pm | 5 | Apr 17 2018 | Assume changes in booths beginning March 18 2020 | [42] |
|  | Su – S: 8am – 8pm | 3 | May 17 2020 |  | [43] |
| Inner City Health  230 Murray St.  Ottawa | 24/7 | 8 | Nov 1 2017 | Assume additional booths added towards end of first year of operation (Aug 2018) | [44, 45] |
|  | 24/7 | 12 | Jul 3 2019 |  | [46] |
|  | 24/7 | 13 | Nov 6 2019 |  | [41] |
|  | 24/7 | 6 | Mar 18 2020 |  | [47] |
|  | 24/7 | 8 | Nov 2020 |  | [47] |
| Somerset West CHC OPS  55 Eccles St.  Ottawa | M – F: 9am – 4 pm | 4 | May 2 2018 |  | [48, 49] |
|  | Su – S: 8:30am – 7:30pm |  | Jun 2019 |  | [50, 51] |
| Path 525 – NorWest Community Centre  525 Simpson St.  Thunder Bay | M – F: 9am – 5pm | 4 | Nov 27 2018 | Assume number of booths planned are number in operation | [52] |
|  | M – S: 10am – 6pm |  | Mar 24 2020 |  | [53, 54] |
| Guelph CHC  150 Duke St. W  Guelph | Su – S: 10am – 5pm | 2 | May 5 2018 | Assume changes in hours beginning March 2020; No mention of fewer booths | [55, 56] |
|  | Su – S: 9am – 4:30pm | 4 | Feb 22 2020 |  | [57] |
|  | M – F: 9am – 5pm |  | April 2020 |  | [58] |
| Sanguen Health Centre  150 Duke St.  Waterloo | Su – S: 9am – 9pm | 2 | Oct 15 2019 |  | [59] |
|  | Su – S: 9am – 9pm | 5 | Oct 9 2020 |  | [60] |

CHC = Community Health Centre; CTS = Consumption and Treatment Service; SIS = Safe injection site; OPS=Overdose Prevention Site; Days of the week: Su=Sunday, M=Monday, T=Tuesday, W=Wednesday, R=Thursday, F=Friday, S=Saturday;

Monthly booth hours per treated public health unit are: $y_{it}=\sum_{n=1}^{N} (b_{nt}\times h_{nt})$

where *y* is the total booth-hours per treated public health unit *i* at month *t*, *b* is the number of booths in operation at a specific site *n*, and *h* is the number of hours operated that month.

Table 2. Naloxone kits dispensed by Public Health Units and community-based organizations

| PHU | 2014 | 2015 | 2016 | 2017 | 2018 | 2019 | 2020 | 2021 | Source |
| --- | --- | --- | --- | --- | --- | --- | --- | --- | --- |
| Algoma |  |  |  | 355 | 590 | 737 |  |  | [61-63] |
| Brant County |  |  |  | 172 | 428 | 1365* | 1282* | 5726* | [64-66] |
| Chatham-Kent |  |  |  | 92 | 381 | 342 | 429 | 46~ | [67-69] |
| Hamilton |  |  | 924 | 3402 | 6412 | 17705 | 16613 | 17046^§^ | [70, 71] |
| Durham Region |  |  |  | 458 🡪 | |  |  |  | [72] |
| Eastern Ontario |  |  |  | 13 🡪 | |  |  |  | [72] |
| Grey-Bruce |  |  |  | 437 | 1337 |  |  | 2434 | [73, 74] |
| Haldimand-Norfolk |  |  |  | 0 | 352 |  |  |  | [72, 75] |
| Haliburton |  |  |  | 297 | | 802 |  |  | [72, 76] |
| Halton Region |  |  |  | 226 🡪 | | 223 | 320 | 418 | [72, 77] |
| Hastings Prince Edward County |  |  | 39 | 166 | 326 | 890 |  |  | [78, 79] |
| Kingston |  |  |  | 456 | 1419 |  |  |  | [80] |
| Lambton |  |  |  | 508 | 700 | 2015 |  |  | [72, 81, 82] |
| Leeds |  |  |  | 408 🡪 | |  |  |  | [72] |
| London | 40 | 98 | 168 | 848 | 2381 | 5289 |  |  | [83] |
| Niagara |  |  |  | 1482 🡪 | | 4761 | 5885 | 7876 | [72] [84] |
| North Bay |  |  |  | 500 | 2000 | 4000 |  |  | [85] |
| Northwestern |  |  |  | 825 | 816 |  |  |  | [86, 87] |
| Ottawa |  |  | 115 | 1300 | 5500 |  |  |  | [88-90] |
| Peel |  |  |  | 366 | 1376 |  |  |  | [91] |
| Peterborough |  |  |  | 833 |  |  |  |  | [72] |
| Porcupine |  |  |  | 224 | 906 | 2656 |  |  | [92, 93] |
| Renfrew |  |  |  | 53 | 197 | 233 |  |  | [94-96] |
| Simcoe Muskoka |  |  | 462 | 1700 | 1620 | 2595 |  |  | [71, 97, 98] |
| Sudbury |  |  |  | 1235 |  | 10010 | 15438* | 24822* | [72, 99, 100] |
| Thunder Bay | 34 | 32 | 35 | 554 | 2022 | 4216 | 6846 |  | [101] |
| Timiskaming |  |  |  | 18 🡪 | |  |  |  | [72] |
| Toronto | 🡨 2000 | |  | 7717 | 42294 | 51577 | 67012 |  | [1, 102-104] |
| Waterloo |  |  | 677 | 4703 | 6782 |  |  |  | [105, 106] |
| Guelph |  |  |  | 581 🡪 | 1419 |  |  |  | [72, 107] |
| Windson-Essex |  |  |  | 1461 🡪 | 3059 | 5378 |  |  | [72, 108] |
| York |  |  |  | 560 🡪 | | 2380 |  |  | [72, 109] |

*Monthly counts available

^§^Quarterly counts available

~Most recent avaliable

🡪 July 1 2017 – June 30 2018

🡨2011 – 2015, inclusive

Table 3. Crude and adjusted segmented regression output for treated/synthetic control unit pairs and aggregate analysis

|  | Emergency department visits per 100,000 population | | | | Hospitalizations per 100,000 population | | | | Deaths per 100,000 population | | | |
| --- | --- | --- | --- | --- | --- | --- | --- | --- | --- | --- | --- | --- |
|  | Crude (95% CI) | P-value | Adj.* (95% CI) | P-value | Crude (95% CI) | P-value | Adj. (95% CI) | P-value | Crude (95% CI) | P-value | Adj. (95% CI) | P-value |
| Toronto |  |  |  |  |  |  |  |  |  |  |  |  |
| Pre-intervention trend, synthetic control (β_1_) | 0.07 (0.03, 0.10) | 0.0001 | 0.08 (0.00, 0.16) | 0.0375 | 0.00 (0.00, 0.01) | 0.1034 | 0.02 (0.00, 0.04) | 0.1067 | 0.01 (0.00, 0.02) | 0.1381 | 0.00 (-0.03, 0.03) | 0.8854 |
| Pre-intervention level difference, treated (β_2_) | 0.23(-0.81, 1.28) | 0.6616 | -1.70 (-5.97, 2.57) | 0.4340 | 0.04 (-0.12, 0.20) | 0.6450 | 0.07 (-0.10, 0.25) | 0.4235 | 0.17 (-0.09, 0.43) | 0.1985 | 0.48 (-0.87, 1.83) | 0.4863 |
| Pre-intervention trend difference, treated (β_3_) | 0.02 (-0.03, 0.07) | 0.4472 | 0.00 (-0.08, 0.07) | 0.9858 | 0.00 (-0.01, 0.01) | 0.5576 | 0.00 (-0.01, 0.01) | 0.7711 | 0.01 (-0.01, 0.02) | 0.2705 | 0.01 (-0.01, 0.03) | 0.2539 |
| Pre-intervention level difference, synthetic control (β_4_) | 0.14 (-0.84, 1.12) | 0.7731 | 0.28 (-0.78, 1.34) | 0.6025 | 0.08 (-0.07, 0.23) | 0.3162 | 0.06 (-0.11, 0.24) | 0.4881 | 0.05 (-0.19, 0.30) | 0.6737 | 0.20 (-0.05, 0.45) | 0.1191 |
| Post-intervention trend difference, synthetic control (β_5_) | 0.00 (-0.05, 0.04) | 0.8690 | -0.02 (-0.07, 0.04) | 0.5650 | -0.01 (-0.01, 0.00) | 0.0227 | -0.01 (-0.03, 0.00) | 0.0639 | 0.01 (0.00, 0.02) | 0.0880 | 0.02 (-0.01, 0.04) | 0.1555 |
| Post-intervention level difference, treated (β_6_) | 0.84 (-0.54, 2.23) | 0.2331 | 1.58 (0.04, 3.12) | 0.0438 | -0.07 (-0.29, 0.14) | 0.5115 | 0.13 (-0.13, 0.38) | 0.3331 | -0.22 (-0.57, 0.13) | 0.2115 | 0.20 (-0.17, 0.57) | 0.2848 |
| Post-intervention trend difference, treated (β_7_) | -0.05 (-0.11, 0.01) | 0.1367 | -0.02 (-0.09, 0.04) | 0.4745 | 0.00 (-0.01, 0.01) | 0.4801 | 0.00 (-0.01, 0.01) | 0.5156 | 0.00 (-0.02, 0.01) | 0.7928 | 0.01 (-0.01, 0.02) | 0.3482 |
| Booth-hours, per hundred (β_8_) |  |  | -0.28 (-0.90, 0.35) | 0.3877 |  |  | -0.02 (-0.13, 0.08) | 0.6308 |  |  | **-0.17 (-0.32, -0.02)** | 0.0259 |
| Individuals dispensed opioid for pain, per hundred (β_9_) |  |  | -0.13 (-0.39, 0.13) | 0.3303 |  |  | 0.00 (-0.04, 0.05) | 0.8628 |  |  | -0.01 (-0.07, 0.05) | 0.7770 |
| Individuals on OAT, per hundred (β_10_) |  |  | -0.43 (-2.03, 1.18) | 0.6036 |  |  | -1.03 (-2.65, 0.59) | 0.2121 |  |  | 0.42 (-1.07, 1.91) | 0.5804 |
| Naloxone kits dispensed, per hundred (β_11_) |  |  | -0.51 (-1.59, 0.56) | 0.3505 |  |  | -0.18 (-0.37, 0.01) | 0.0573 |  |  | **-0.34 (-0.61, -0.08)** | 0.0113 |
| Hamilton |  |  |  |  |  |  |  |  |  |  |  |  |
| Pre-intervention trend, synthetic control (β_1_) | 0.15 (0.08, 0.21) | 0.0000 | 0.04 (-0.15, 0.22) | 0.6802 | 0.00 (-0.01, 0.02) | 0.6690 | 0.01 (-0.03, 0.06) | 0.5453 | 0.02 (0.00, 0.04) | 0.0177 | 0.01 (-0.04, 0.06) | 0.6947 |
| Pre-intervention level difference, treated (β_2_) | 0.31 (-1.65, 2.27) | 0.7555 | 2.94 (-2.62, 8.49) | 0.2998 | 0.19 (-0.35, 0.72) | 0.4938 | 0.12 (-1.26, 1.51) | 0.8601 | 0.40 (-0.10, 0.91) | 0.1177 | 0.82 (-3.06, 4.66) | 0.6844 |
| Pre-intervention trend difference, treated (β_3_) | 0.00 (-0.09, 0.10) | 0.9469 | 0.02 (-0.08, 0.11) | 0.7488 | 0.00 (-0.03, 0.02) | 0.8184 | 0.00 (-0.03, 0.03) | 0.9570 | 0.01 (-0.01, 0.04) | 0.2362 | 0.02 (-0.01, 0.04) | 0.2771 |
| Pre-intervention level difference, synthetic control (β_4_) | 0.28 (-1.66, 2.22) | 0.7770 | 0.68 (-1.41, 2.77) | 0.5231 | -0.27 (-0.80, 0.25) | 0.3075 | -0.19 (-0.77, 0.39) | 0.5135 | -0.12 (-0.62, 0.38) | 0.6430 | -0.13 (-0.70, 0.43) | 0.6448 |
| Post-intervention trend difference, synthetic control (β_5_) | -0.09 (-0.19, 0.01) | 0.0662 | 0.02 (-0.16, 0.20) | 0.8288 | 0.00 (-0.02, 0.03) | 0.7966 | 0.00 (-0.02, 0.03) | 0.7751 | 0.05 (0.02, 0.07) | 0.0003 | 0.05 (0.01, 0.09) | 0.0164 |
| Post-intervention level difference, treated (β_6_) | 1.31 (-1.44, 4.06) | 0.3499 | 7.68 (-0.63, 16.00) | 0.0700 | 0.00 (-0.74, 0.75) | 0.9929 | 0.13 (-2.21, 2.46) | 0.9163 | -0.06 (-0.77, 0.65) | 0.8697 | **2.98 (0.79, 5.16)** | **0.0075** |
| Post-intervention trend difference, treated (β_7_) | -0.05 (-0.18, 0.09) | 0.4998 | -0.20 (-0.40, 0.01) | 0.0603 | 0.01 (-0.03, 0.05) | 0.6476 | 0.00 (-0.08, 0.08) | 0.9995 | **-0.07 (-0.10, -0.03)** | **0.0002** | **-0.12 (-0.17, -0.06)** | **<0.0001** |
| Booth-hours, per hundred (β_8_) |  |  | -3.60 (-8.25, 1.06) | 0.1297 |  |  | -0.06 (-1.41, 1.28) | 0.9250 |  |  | **-1.68 (-2.87, -0.48)** | 0.0060 |
| Individuals dispensed opioid for pain, per hundred (β_9_) |  |  | 0.12 (-0.25, 0.48) | 0.5262 |  |  | 0.04 (-0.06, 0.15) | 0.4270 |  |  | 0.00 (-0.10, 0.09) | 0.9323 |
| Individuals on OAT, per hundred (β_10_) |  |  | 2.59 (-2.55, 7.74) | 0.3237 |  |  | -0.15 (-1.62, 1.33) | 0.8467 |  |  | 0.13 (-1.05, 1.31) | 0.8314 |
| Naloxone kits dispensed, per hundred (β_11_) |  |  | 0.86 (-0.11, 1.84) | 0.0827 |  |  | -0.06 (-0.35, 0.23) | 0.6738 |  |  | 0.07 (-0.12, 0.27) | 0.4566 |
| Kingston |  |  |  |  |  |  |  |  |  |  |  |  |
| Pre-intervention trend, synthetic control (β_1_) | 0.15 (0.07, 0.23) | 0.0002 | 0.15 (-0.05, 0.35) | 0.1405 | 0.02 (0.00, 0.05) | 0.0713 | 0.08 (0.00, 0.16) | 0.0427 | 0.01 (0.00, 0.03) | 0.1407 | -0.01 (-0.07, 0.04) | 0.5804 |
| Pre-intervention level difference, treated (β_2_) | 1.23 (-1.08, 3.54) | 0.2964 | 1.42 (-6.06, 8.90) | 0.7098 | 0.37 (-0.40, 1.15) | 0.3462 | -2.52 (-6.25, 1.21) | 0.1847 | 0.10 (0.00, 0.03) | 0.7204 | 0.85 (-0.39, 2.08) | 0.1788 |
| Pre-intervention trend difference, treated (β_3_) | 0.07 (-0.04, 0.18) | 0.2292 | 0.12 (0.00, 0.25) | 0.0510 | 0.02 (-0.02, 0.06) | 0.2594 | 0.01 (-0.04, 0.05) | 0.8403 | 0.00 (-0.02, 0.03) | 0.8128 | 0.02 (-0.02, 0.05) | 0.3146 |
| Pre-intervention level difference, synthetic control (β_4_) | -0.28 (-2.58, 2.03) | 0.8141 | 0.11 (-2.81, 3.02) | 0.9432 | -0.73 (-1.51, 0.04) | 0.0629 | -1.16 (-2.05, -0.27) | 0.0104 | -0.09 (-1.04, 0.86) | 0.7562 | -0.09 (-0.69, 0.51) | 0.7654 |
| Post-intervention trend difference, synthetic control (β_5_) | 0.04 (-0.08, 0.15) | 0.5192 | 0.04 (-0.11, 0.18) | 0.6327 | -0.02 (-0.05, 0.02) | 0.4486 | -0.05 (-0.10, 0.00) | 0.0566 | 0.02 (0.00, 0.05) | 0.0875 | 0.04 (0.00, 0.07) | 0.0406 |
| Post-intervention level difference, treated (β_6_) | -1.51 (-4.77, 1.76) | 0.3655 | -6.03 (-16.05, 3.98) | 0.2376 | 0.76 (-0.34, 1.85) | 0.1757 | 2.22 (-1.08, 5.52) | 0.1869 | 0.23 (-0.55, 1.01) | 0.5606 | -0.98 (-3.26, 1.30) | 0.4000 |
| Post-intervention trend difference, treated (β_7_) | -0.13 (-0.29, 0.04) | 0.1257 | -0.19 (-0.40, 0.02) | 0.0721 | **-0.07 (-0.13, -0.02)** | 0.0118 | -0.04 (-0.11, 0.03) | 0.2177 | -0.03 (-0.07, 0.01) | 0.1277 | -0.05 (-0.10, 0.00) | 0.0654 |
| Booth-hours, per hundred (β_8_) |  |  | 1.41 (-1.46, 4.29) | 0.3356 |  |  | -0.39 (-1.36, 0.57) | 0.4237 |  |  | 0.37 (-0.31, 1.05) | 0.2821 |
| Individuals dispensed opioid for pain, per hundred (β_9_) |  |  | **0.62 (0.12, 1.11)** | **0.0144** |  |  | **0.16 (0.00, 0.32)** | **0.0449** |  |  | 0.02 (-0.10, 0.14) | 0.7855 |
| Individuals on OAT, per hundred (β_10_) |  |  | 0.19 (-5.91, 6.29) | 0.9518 |  |  | -1.86 (-3.95, 0.24) | 0.0820 |  |  | 0.52 (-0.27, 1.30) | 0.1979 |
| Naloxone kits dispensed, per hundred (β_11_) |  |  | 1.12 (-0.21, 2.46) | 0.0997 |  |  | 0.37 (-0.07, 0.81) | 0.1033 |  |  | 0.19 (-0.25, 0.64) | 0.3972 |
| London |  |  |  |  |  |  |  |  |  |  |  |  |
| Pre-intervention trend, synthetic control (β_1_) | 0.10 (0.02, 0.18) | 0.0120 | 0.23 (0.05, 0.41) | 0.0114 | 0.00 (-0.02, 0.02) | 0.8755 | 0.03 (-0.01, 0.06) | 0.1078 | 0.01 (-0.01, 0.02) | 0.2685 | 0.05 (0.01, 0.08) | 0.0076 |
| Pre-intervention level difference, treated (β_2_) | 0.78 (-1.57, 3.12) | 0.5164 | -1.25 (-7.23, 4.74) | 0.6834 | 0.16 (-0.37, 0.69) | 0.5634 | 1.12 (0.00, 2.24) | 0.0492 | 0.15 (-0.25, 0.55) | 0.4642 | 1.52 (0.41, 2.63) | 0.0072 |
| Pre-intervention trend difference, treated (β_3_) | 0.04 (-0.07, 0.15) | 0.5191 | -0.03 (-0.18, 0.13) | 0.7339 | 0.00 (-0.02, 0.03) | 0.7280 | 0.01 (-0.02, 0.03) | 0.5730 | 0.00 (-0.01, 0.02) | 0.6699 | -0.01 (-0.03, 0.01) | 0.4646 |
| Pre-intervention level difference, synthetic control (β_4_) | -0.64 (-2.90, 1.63) | 0.5816 | -0.62 (-2.87, 1.63) | 0.5883 | -0.47 (-0.98, 0.05) | 0.0747 | -0.25 (-0.77, 0.28) | 0.3583 | -0.12 (-0.51, 0.27) | 0.5341 | -0.05 (-0.45, 0.35) | 0.8049 |
| Post-intervention trend difference, synthetic control (β_5_) | 0.21 (0.10, 0.32) | 0.0001 | **0.23 (0.12, 0.35)** | 0.0001 | 0.01 (-0.01, 0.04) | 0.2402 | 0.01 (-0.02, 0.04) | 0.5853 | 0.02 (0.00, 0.04) | 0.0573 | 0.00 (-0.03, 0.03) | 0.9849 |
| Post-intervention level difference, treated (β_6_) | 0.61 (-2.60, 3.81) | 0.7108 | **5.34 (1.61, 9.08)** | 0.0050 | 0.50 (-0.22, 1.23) | 0.1755 | **1.06 (0.18, 1.95)** | 0.0186 | -0.01 (-0.56, 0.54) | 0.9719 | 0.51 (-0.15, 1.17) | 0.1330 |
| Post-intervention trend difference, treated (β_7_) | -0.13 (-0.28, 0.02) | 0.0949 | **-0.19 (-0.37, -0.01)** | 0.0413 | **-0.04 (-0.07, 0.00)** | 0.0271 | **-0.06 (-0.11, -0.02)** | 0.0107 | 0.01 (-0.02, 0.03) | 0.5354 | -0.02 (-0.06, 0.02) | 0.3069 |
| Booth-hours, per hundred (β_8_) |  |  | **-2.97 (-4.45, -1.49)** | 0.0001 |  |  | -0.37 (-0.77, 0.02) | 0.0659 |  |  | **-0.45 (-0.76, -0.14)** | 0.0041 |
| Individuals dispensed opioid for pain, per hundred (β_9_) |  |  | 0.37 (-0.05, 0.79) | 0.0832 |  |  | 0.08 (-0.01, 0.18) | 0.0923 |  |  | 0.06 (-0.01, 0.13) | 0.0890 |
| Individuals on OAT, per hundred (β_10_) |  |  | -1.69 (-4.41, 1.03) | 0.2238 |  |  | -0.85 (-2.33, 0.63) | 0.2614 |  |  | **-1.28 (-2.49, -0.07)** | 0.0389 |
| Naloxone kits dispensed, per hundred (β_11_) |  |  | -0.22 (-0.99, 0.56) | 0.5855 |  |  | **-0.34 (-0.56, -0.12)** | 0.0023 |  |  | -0.11 (-0.28, 0.06) | 0.1867 |

|  | Emergency department visits per 100,000 population | | | | Hospitalizations per 100,000 population | | | | Deaths per 100,000 population | | | |
| --- | --- | --- | --- | --- | --- | --- | --- | --- | --- | --- | --- | --- |
|  | Crude (95% CI) | P-value | Adj.* (95% CI) | P-value | Crude (95% CI) | P-value | Adj. (95% CI) | P-value | Crude (95% CI) | P-value | Adj. (95% CI) | P-value |
| Niagara |  |  |  |  |  |  |  |  |  |  |  |  |
| Pre-intervention trend, synthetic control (β_1_) | 0.17 (0.07, 0.27) | 0.0008 | 0.16 (-0.08, 0.40) | 0.2022 | 0.00 (-0.02, 0.02) | 0.8813 | 0.00 (-0.05, 0.05) | 0.9961 | 0.03 (0.01, 0.05) | 0.0010 | 0.01 (-0.03, 0.05) | 0.6682 |
| Pre-intervention level difference, treated (β_2_) | 4.52 (1.49, 7.54) | 0.0034 | 3.73 (0.76, 6.70) | 0.0138 | 0.45 (-0.16, 1.07) | 0.1506 | 0.09 (-0.70, 0.89) | 0.8185 | 0.43 (-0.11, 0.97) | 0.1198 | 0.52 (-0.10, 1.14) | 0.0983 |
| Pre-intervention trend difference, treated (β_3_) | 0.13 (-0.01, 0.27) | 0.0698 | 0.13 (-0.01, 0.27) | 0.0673 | 0.02 (-0.01, 0.05) | 0.1480 | 0.03 (0.00, 0.06) | 0.0648 | 0.01 (-0.02, 0.03) | 0.6772 | 0.00 (-0.02, 0.03) | 0.8667 |
| Pre-intervention level difference, synthetic control (β_4_) | 0.31 (-2.84, 3.46) | 0.8466 | -0.20 (-3.51, 3.12) | 0.9076 | -0.84 (-1.48, -0.20) | 0.0106 | -0.65 (-1.36, 0.07) | 0.0754 | 0.13 (-0.43, 0.70) | 0.6462 | 0.00 (-0.64, 0.65) | 0.9982 |
| Post-intervention trend difference, synthetic control (β_5_) | -0.12 (-0.30, 0.06) | 0.1992 | -0.08 (-0.30, 0.14) | 0.4772 | 0.02 (-0.01, 0.06) | 0.2387 | 0.03 (-0.02, 0.07) | 0.2670 | 0.00 (-0.03, 0.03) | 0.8787 | 0.02 (-0.02, 0.06) | 0.4082 |
| Post-intervention level difference, treated (β_6_) | -2.65 (-7.10, 1.81) | 0.2440 | **25.88 (6.44, 45.33)** | 0.0091 | 0.30 (-0.61, 1.21) | 0.5241 | 2.92 (-1.32, 7.14) | 0.1775 | -0.67 (-1.47, 0.13) | 0.0983 | 4.67 (0.99, 8.35) | 0.0129 |
| Post-intervention trend difference, treated (β_7_) | -0.13 (-0.38, 0.12) | 0.3130 | **-0.49 (-0.82, -0.15)** | 0.0044 | -0.05 (-0.10, 0.00) | 0.0508 | **-0.08 (-0.16, -0.01)** | 0.0219 | 0.03 (-0.01, 0.08) | 0.1364 | -0.03 (-0.09, 0.04) | 0.3798 |
| Booth-hours, per hundred (β_8_) |  |  | **-9.70 (-16.08, -3.31)** | 0.0029 |  |  | -0.89 (-2.28, 0.50) | 0.2111 |  |  | **-1.78 (-2.99, -0.58)** | 0.0037 |
| Individuals dispensed opioid for pain, per hundred (β_9_) |  |  | 0.40 (-0.16, 0.97) | 0.1583 |  |  | 0.07 (-0.05, 0.19) | 0.2614 |  |  | -0.07 (-0.17, 0.04) | 0.2103 |
| Individuals on OAT, per hundred (β_10_) |  |  | -2.35 (-9.09, 4.38) | 0.4934 |  |  | 0.63 (-0.90, 2.17) | 0.4205 |  |  | 0.23 (-1.06, 1.52) | 0.7302 |
| Naloxone kits dispensed, per hundred (β_11_) |  |  | **2.49 (0.98, 4.00)** | 0.0013 |  |  | 0.02 (-0.33, 0.36) | 0.9290 |  |  | 0.11 (-0.18, 0.39) | 0.4581 |
| Ottawa |  |  |  |  |  |  |  |  |  |  |  |  |
| Pre-intervention trend, synthetic control (β_1_) | 0.05 (0.02, 0.08) | 0.0011 | 0.03 (-0.03, 0.09) | 0.3768 | 0.01 (0.00, 0.01) | 0.079 | 0.00 (-0.02, 0.02) | 0.6788 | 0.00 (0.00, 0.01) | 0.3272 | 0.01 (-0.01, 0.03) | 0.2387 |
| Pre-intervention level difference, treated (β_2_) | 0.03 (-0.83, 0.89) | 0.9450 | 0.30 (-2.86, 3.46) | 0.8524 | 0.07 (-0.16, 0.29) | 0.5515 | 0.31 (-0.53, 1.15) | 0.4679 | 0.03 (-0.18, 0.25) | 0.7589 | -0.26 (-1.01, 0.49) | 0.4997 |
| Pre-intervention trend difference, treated (β_3_) | 0.00 (-0.04, 0.04) | 0.9889 | -0.01 (-0.04, 0.03) | 0.7889 | 0.00 (-0.01, 0.01) | 0.8868 | 0.00 (-0.01, 0.02) | 0.8182 | 0.00 (-0.01, 0.01) | 0.6302 | 0.00 (-0.01, 0.01) | 0.8024 |
| Pre-intervention level difference, synthetic control (β_4_) | 0.84 (0.03, 1.64) | 0.0425 | 0.70 (-0.03, 1.42) | 0.0590 | -0.03 (-0.24, 0.18) | 0.8132 | -0.02 (-0.25, 0.22) | 0.8926 | 0.04 (-0.16, 0.25) | 0.6672 | 0.01 (-0.19, 0.22) | 0.9060 |
| Post-intervention trend difference, synthetic control (β_5_) | -0.03 (-0.06, 0.01) | 0.1357 | -0.02 (-0.08, 0.04) | 0.4980 | -0.01 (-0.02, 0.00) | 0.0417 | -0.01 (-0.02, 0.01) | 0.3411 | 0.01 (0.00, 0.02) | 0.1311 | 0.00 (-0.01, 0.02) | 0.8950 |
| Post-intervention level difference, treated (β_6_) | **-1.32 (-2.47, -0.18)** | 0.0233 | 0.62 (-0.59, 1.84) | 0.3155 | -0.24 (-0.54, 0.05) | 0.1087 | -0.23 (-0.53, 0.12) | 0.1911 | -0.06 (-0.35, 0.23) | 0.6831 | 0.27 (-0.06, 0.60) | 0.1094 |
| Post-intervention trend difference, treated (β_7_) | **0.07 (0.02, 0.12)** | 0.0057 | **0.05 (0.00, 0.11)** | 0.0381 | 0.00 (-0.01, 0.02) | 0.7004 | 0.00 (-0.01, 0.02) | 0.9773 | 0.00 (-0.01, 0.01) | 0.8765 | 0.01 (-0.01, 0.02) | 0.4505 |
| Booth-hours, per hundred (β_8_) |  |  | **-0.26 (-0.34, -0.17)** | **<0.0001** |  |  | 0.00 (-0.03, 0.02) | 0.7200 |  |  | **-0.04 (-0.07, -0.02)** | 0.0008 |
| Individuals dispensed opioid for pain, per hundred (β_9_) |  |  | -0.12 (-0.30, 0.07) | 0.2203 |  |  | 0.04 (-0.01, 0.09) | 0.1559 |  |  | -0.01 (-0.06, 0.05) | 0.8074 |
| Individuals on OAT, per hundred (β_10_) |  |  | 1.09 (-4.38, 6.56) | 0.6967 |  |  | 0.24 (-0.68, 1.16) | 0.6078 |  |  | -0.60 (-2.02, 0.82) | 0.4099 |
| Naloxone kits dispensed, per hundred (β_11_) |  |  | **0.40 (0.03, 0.78)** | 0.0354 |  |  | 0.05 (-0.06, 0.16) | 0.3325 |  |  | -0.03 (-0.13, 0.08) | 0.6314 |
| Thunder Bay |  |  |  |  |  |  |  |  |  |  |  |  |
| Pre-intervention trend, synthetic control (β_1_) | 0.15 (0.07, 0.24) | 0.0005 | 0.10 (-0.03, 0.23) | 0.1237 | 0.01 (-0.02, 0.04) | 0.3679 | 0.01 (-0.03, 0.06) | 0.5316 | 0.03 (0.00, 0.06) | 0.0716 | 0.05 (0.00, 0.10) | 0.0306 |
| Pre-intervention level difference, treated (β_2_) | 3.59 (1.00, 6.18) | 0.0067 | -1.44 (-15.19, 12.31) | 0.8374 | 0.61 (-0.26, 1.47) | 0.1689 | 1.77 (-2.68, 6.22) | 0.4355 | 1.22 (0.30, 2.13) | 0.0095 | 3.15 (-1.26, 7.55) | 0.1613 |
| Pre-intervention trend difference, treated (β_3_) | 0.12 (0.00, 0.24) | 0.0556 | 0.10 (-0.04, 0.24) | 0.1531 | 0.02 (-0.02, 0.06) | 0.4362 | 0.02 (-0.03, 0.06) | 0.4571 | 0.03 (-0.01, 0.08) | 0.1346 | 0.04 (0.00, 0.09) | 0.0676 |
| Pre-intervention level difference, synthetic control (β_4_) | -0.31 (-2.99, 2.37) | 0.8210 | -0.36 (-3.19, 2.46) | 0.8005 | -0.55 (-1.44, 0.35) | 0.2307 | -0.54 (-1.50, 0.42) | 0.2690 | -0.10 (-1.04, 0.85) | 0.8431 | -0.12 (-1.13, 0.88) | 0.8104 |
| Post-intervention trend difference, synthetic control (β_5_) | 0.31 (0.16, 0.46) | 0.0000 | 0.28 (0.13, 0.43) | 0.0002 | 0.00 (-0.05, 0.05) | 0.8802 | 0.00 (-0.05, 0.05) | 0.8938 | 0.04 (-0.02, 0.09) | 0.1884 | 0.03 (-0.02, 0.08) | 0.2482 |
| Post-intervention level difference, treated (β_6_) | -1.75 (-5.54, 2.03) | 0.3640 | -0.10 (-6.73, 6.53) | 0.9758 | -0.49 (-1.72, 0.81) | 0.4825 | -0.65 (-2.99, 1.69) | 0.5855 | -0.86 (-2.20, 0.48) | 0.2087 | 1.26 (-1.13, 3.65) | 0.3021 |
| Post-intervention trend difference, treated (β_7_) | **-0.36 (-0.57, -0.15)** | 0.0007 | **-0.35 (-0.60, -0.10)** | 0.0054 | -0.01 (-0.08, 0.06) | 0.6805 | -0.01 (-0.10, 0.07) | 0.7742 | 0.00 (-0.08, 0.07) | 0.9259 | 0.05 (-0.04, 0.13) | 0.2837 |
| Booth-hours, per hundred (β_8_) |  |  | -0.43 (-1.89, 1.03) | 0.5630 |  |  | 0.03 (-0.48, 0.54) | 0.9065 |  |  | **-0.55 (-1.08, -0.03)** | **0.0398** |
| Individuals dispensed opioid for pain, per hundred (β_9_) |  |  | 0.27 (-0.13, 0.68) | 0.1805 |  |  | 0.02 (-0.13, 0.17) | 0.7678 |  |  | 0.01 (-0.15, 0.17) | 0.8994 |
| Individuals on OAT, per hundred (β_10_) |  |  | 0.52 (-0.95, 1.99) | 0.4879 |  |  | -0.14 (-0.62, 0.33) | 0.5619 |  |  | -0.21 (-0.69, 0.27) | 0.3947 |
| Naloxone kits dispensed, per hundred (β_11_) |  |  | **0.85 (0.05, 1.66)** | 0.0378 |  |  | 0.14 (-0.19, 0.47) | 0.4155 |  |  | -0.15 (-0.45, 0.16) | 0.3416 |
| Guelph |  |  |  |  |  |  |  |  |  |  |  |  |
| Pre-intervention trend, synthetic control (β_1_) | 0.07 (0.03, 0.12) | 0.0016 | 0.09 (-0.02, 0.21) | 0.1035 | 0.01 (-0.01, 0.03) | 0.2880 | 0.00 (-0.05, 0.05) | 0.9854 | 0.01 (0.00, 0.03) | 0.0815 | 0.02 (-0.02, 0.06) | 0.3150 |
| Pre-intervention level difference, treated (β_2_) | 0.41 (-0.96, 1.77) | 0.5579 | -1.02 (-6.45, 4.42) | 0.7140 | -0.08 (-0.61, 0.45) | 0.7752 | -0.10 (-2.56, 2.37) | 0.9395 | -0.08 (-0.50, 0.35) | 0.7263 | -0.16 (-2.26, 1.94) | 0.8794 |
| Pre-intervention trend difference, treated (β_3_) | 0.01 (-0.06, 0.07) | 0.8003 | -0.01 (-0.11, 0.08) | 0.8096 | 0.00 (-0.03, 0.02) | 0.9755 | 0.00 (-0.04, 0.04) | 0.9861 | 0.00 (-0.02, 0.02) | 0.9159 | 0.00 (-0.03, 0.03) | 0.8596 |
| Pre-intervention level difference, synthetic control (β_4_) | 0.48 (-0.86, 1.83) | 0.4812 | 0.36 (-1.01, 1.74) | 0.6020 | -0.18 (-0.70, 0.34) | 0.4952 | -0.19 (-0.75, 0.38) | 0.5170 | -0.03 (-0.45, 0.39) | 0.8806 | 0.05 (-0.50, 0.60) | 0.8598 |
| Post-intervention trend difference, synthetic control (β_5_) | -0.06 (-0.12, 0.01) | 0.0992 | -0.07 (-0.17, 0.03) | 0.1802 | -0.01 (-0.03, 0.02) | 0.5010 | -0.01 (-0.05, 0.02) | 0.5221 | 0.01 (-0.01, 0.03) | 0.4019 | 0.01 (-0.02, 0.04) | 0.6366 |
| Post-intervention level difference, treated (β_6_) | 1.38 (-0.52, 3.29) | 0.1535 | -1.13 (-3.87, 1.61) | 0.4198 | 0.40 (-0.34, 1.14) | 0.2890 | 0.07 (-1.06, 1.19) | 0.9092 | 0.29 (-0.31, 0.88) | 0.3491 | 0.04 (-0.96, 1.04) | 0.9392 |
| Post-intervention trend difference, treated (β_7_) | -0.07 (-0.16, 0.03) | 0.1655 | -0.16 (-0.38, 0.06) | 0.1519 | -0.02 (-0.06, 0.01) | 0.2232 | -0.04 (-0.12, 0.04) | 0.2926 | -0.02 (-0.05, 0.01) | 0.1789 | -0.01 (-0.08, 0.05) | 0.7275 |
| Booth-hours, per hundred (β_8_) |  |  | **2.39 (0.73, 4.04)** | 0.0046 |  |  | 0.28 (-0.40, 0.95) | 0.4194 |  |  | 0.17 (-0.36, 0.71) | 0.5265 |
| Individuals dispensed opioid for pain, per hundred (β_9_) |  |  | 0.01 (-0.29, 0.32) | 0.9394 |  |  | -0.07 (-0.20, 0.05) | 0.2549 |  |  | 0.04 (-0.06, 0.14) | 0.4112 |
| Individuals on OAT, per hundred (β_10_) |  |  | -1.31 (-6.12, 3.50) | 0.5923 |  |  | 0.02 (-2.03, 2.07) | 0.9873 |  |  | -0.09 (-1.73, 1.55) | 0.9161 |
| Naloxone kits dispensed, per hundred (β_11_) |  |  | 0.18 (-0.74, 1.10) | 0.7055 |  |  | 0.06 (-0.30, 0.42) | 0.7386 |  |  | -0.07 (-0.33, 0.19) | 0.5955 |

|  | Emergency department visits per 100,000 population | | | | Hospitalizations per 100,000 population | | | | Deaths per 100,000 population | | | |
| --- | --- | --- | --- | --- | --- | --- | --- | --- | --- | --- | --- | --- |
|  | Crude (95% CI) | P-value | Adj.* (95% CI) | P-value | Crude (95% CI) | P-value | Adj. (95% CI) | P-value | Crude (95% CI) | P-value | Adj. (95% CI) | P-value |
| Waterloo |  |  |  |  |  |  |  |  |  |  |  |  |
| Pre-intervention trend, synthetic control (β_1_) | 0.13 (0.07, 0.18) | 0.0000 | 0.00 (-0.16, 0.16) | 0.9817 | 0.00 (-0.01, 0.01) | 0.8837 | -0.05 (-0.08, -0.02) | 0.0023 | 0.02 (0.00, 0.03) | 0.0308 | -0.01 (-0.05, 0.04) | 0.7641 |
| Pre-intervention level difference, treated (β_2_) | -0.56 (-2.24, 1.11) | 0.5111 | 9.39 (-3.57, 22.35) | 0.1555 | -0.26 (-0.66, 0.14) | 0.1971 | 1.90 (-0.38, 4.17) | 0.1027 | -0.15 (-0.61, 0.30) | 0.5053 | 0.52 (-1.90, 2.93) | 0.6737 |
| Pre-intervention trend difference, treated (β_3_) | -0.04 (-0.12, 0.04) | 0.2936 | -0.01 (-0.10, 0.08) | 0.8676 | -0.01 (-0.03, 0.01) | 0.1961 | -0.02 (-0.04, 0.00) | 0.0438 | -0.02 (-0.04, 0.01) | 0.1493 | -0.01 (-0.04, 0.02) | 0.4254 |
| Pre-intervention level difference, synthetic control (β_4_) | -1.52 (-3.49, 0.46) | 0.1319 | -0.72 (-2.83, 1.40) | 0.5072 | -0.41 (-0.88, 0.06) | 0.0837 | -0.44 (-0.92, 0.05) | 0.0766 | -0.37 (-0.90, 0.16) | 0.1736 | -0.21 (-0.82, 0.39) | 0.4918 |
| Post-intervention trend difference, synthetic control (β_5_) | 0.25 (0.09, 0.42) | 0.0031 | 0.19 (0.01, 0.37) | 0.0423 | 0.04 (0.00, 0.08) | 0.0460 | 0.05 (0.01, 0.09) | 0.0088 | 0.07 (0.02, 0.11) | 0.0036 | 0.07 (0.01, 0.13) | 0.0142 |
| Post-intervention level difference, treated (β_6_) | -0.38 (-3.17, 2.41) | 0.7876 | -1.20 (-4.09, 1.70) | 0.4187 | 0.69 (0.03, 1.35) | 0.0415 | **0.73 (0.05, 1.54)** | **0.0359** | 0.16 (-0.59, 0.92) | 0.6696 | 0.03 (-0.81, 0.88) | 0.9373 |
| Post-intervention trend difference, treated (β_7_) | -0.21 (-0.45, 0.02) | 0.0775 | -0.05 (-0.46, 0.36) | 0.8119 | -0.03 (-0.08, 0.03) | 0.3645 | 0.05 (-0.05, 0.15) | 0.2914 | -0.01 (-0.08, 0.05) | 0.6739 | 0.01 (-0.11, 0.13) | 0.8911 |
| Booth-hours, per hundred (β_8_) |  |  | 0.21 (-1.48, 1.91) | 0.8065 |  |  | -0.14 (-0.54, 0.27) | 0.5092 |  |  | -0.01 (-0.50, 0.48) | 0.9687 |
| Individuals dispensed opioid for pain, per hundred (β_9_) |  |  | -0.09 (-0.53, 0.34) | 0.6705 |  |  | **-0.22 (-0.33, -0.10)** | **0.0002** |  |  | -0.02 (-0.15, 0.11) | 0.7523 |
| Individuals on OAT, per hundred (β_10_) |  |  | 5.05 (-1.72, 11.82) | 0.1437 |  |  | **2.62 (0.02, 5.22)** | **0.0480** |  |  | 0.64 (-1.52, 2.80) | 0.5617 |
| Naloxone kits dispensed, per hundred (β_11_) |  |  | 0.77 (-0.16, 1.71) | 0.1052 |  |  | 0.18 (-0.08, 0.45) | 0.1758 |  |  | 0.20 (-0.11, 0.52) | 0.2042 |
| Aggregate |  |  |  |  |  |  |  |  |  |  |  |  |
| Pre-intervention trend, synthetic control (β_1_) | 0.08 (0.06, 0.09) | <0.0001 | 0.18 (0.09, 0.26) | 0.0001 | 0.00 (0.00, 0.00) | 0.0190 | 0.02 (0.00, 0.04) | 0.0830 | 0.01 (0.00, 0.01) | <0.0001 | 0.06 (0.03, 0.08) | <0.0001 |
| Pre-intervention level difference, treated (β_2_) | 1.63 (1.21, 2.06) | <0.0001 | 3.01 (1.81, 4.20) | <0.0001 | 0.33 (0.22, 0.44) | <0.0001 | 1.22 (-0.37, 2.82) | 0.1332 | 0.33 (0.23, 0.44) | <0.0001 | 1.45 (0.91, 1.99) | <0.0001 |
| Pre-intervention trend difference, treated (β_3_) | 0.05 (0.03, 0.07) | <0.0001 | 0.02 (0.01, 0.04) | 0.0101 | 0.00 (0.00, 0.01) | 0.1387 | 0.01 (0.00, 0.02) | 0.1106 | 0.01 (0.00, 0.01) | 0.0015 | 0.01 (0.00, 0.01) | 0.0090 |
| Pre-intervention level difference, synthetic control (β_4_) | 0.59 (0.03, 1.15) | 0.0395 | 0.08 (-0.41, 0.57) | 0.7569 | -0.06 (-0.21, 0.09) | 0.4142 | -0.09 (-0.25, 0.07) | 0.2649 | 0.09 (-0.05, 0.23) | 0.2032 | 0.01 (-0.13, 0.14) | 0.9375 |
| Post-intervention trend difference, synthetic control (β_5_) | -0.04 (-0.09, 0.01) | 0.0857 | -0.15 (-0.22, -0.09) | <0.0001 | -0.01 (-0.02, 0.00) | 0.2137 | -0.02 (-0.03, 0.00) | 0.0621 | 0.00 (-0.01, 0.02) | 0.6299 | -0.03 (-0.05, -0.01) | 0.0013 |
| Post-intervention level difference, treated (β_6_) | -0.33 (-1.12, 0.46) | 0.4041 | **-0.79 (-1.54, -0.04)** | 0.0400 | 0.05 (-0.16, 0.26) | 0.6173 | 0.03 (-0.22, 0.27) | 0.8392 | -0.10 (-0.30, 0.10) | 0.3205 | -0.17 (-0.38, 0.04) | 0.1143 |
| Post-intervention trend difference, treated (β_7_) | -0.07 (-0.14, 0.00) | 0.0517 | **-0.15 (-0.21, -0.08)** | <0.0001 | -0.02 (-0.03, 0.00) | 0.0685 | **-0.02 (-0.05, 0.00)** | 0.0269 | -0.01 (-0.02, 0.01) | 0.3967 | -0.01 (-0.03, 0.01) | 0.4378 |
| Booth-hours, per hundred (β_8_) |  |  | 0.19 (-0.02, 0.40) | 0.0760 |  |  | 0.05 (-0.03, 0.13) | 0.6638 |  |  | 0.05 (-0.01, 0.11) | 0.0780 |
| Individuals dispensed opioid for pain, per hundred (β_9_) |  |  | 0.04 (-0.19, 0.27) | 0.7469 |  |  | -1.21 (-3.00, 0.58) | 0.2058 |  |  | 0.03 (-0.03, 0.10) | 0.3187 |
| Individuals on OAT, per hundred (β_10_) |  |  | **-9.05 (-14.16, -3.94)** | 0.0005 |  |  | -1.21 (-3.00, 0.58) | 0.1861 |  |  | **-3.35 (-4.89, -1.81)** | <0.0001 |
| Naloxone kits dispensed, per hundred (β_11_) |  |  | 0.73 (-0.31, 1.76) | 0.1694 |  |  | -0.05 (-0.39, 0.30) | 0.7848 |  |  | **-0.38 (-0.67, -0.08)** | 0.0119 |

*Adjusted for seasonality using harmonic terms and with robust standard errors generated using Newey-West method, except for aggregate analysis which does not adjust for seasonality given multiple baseline approach

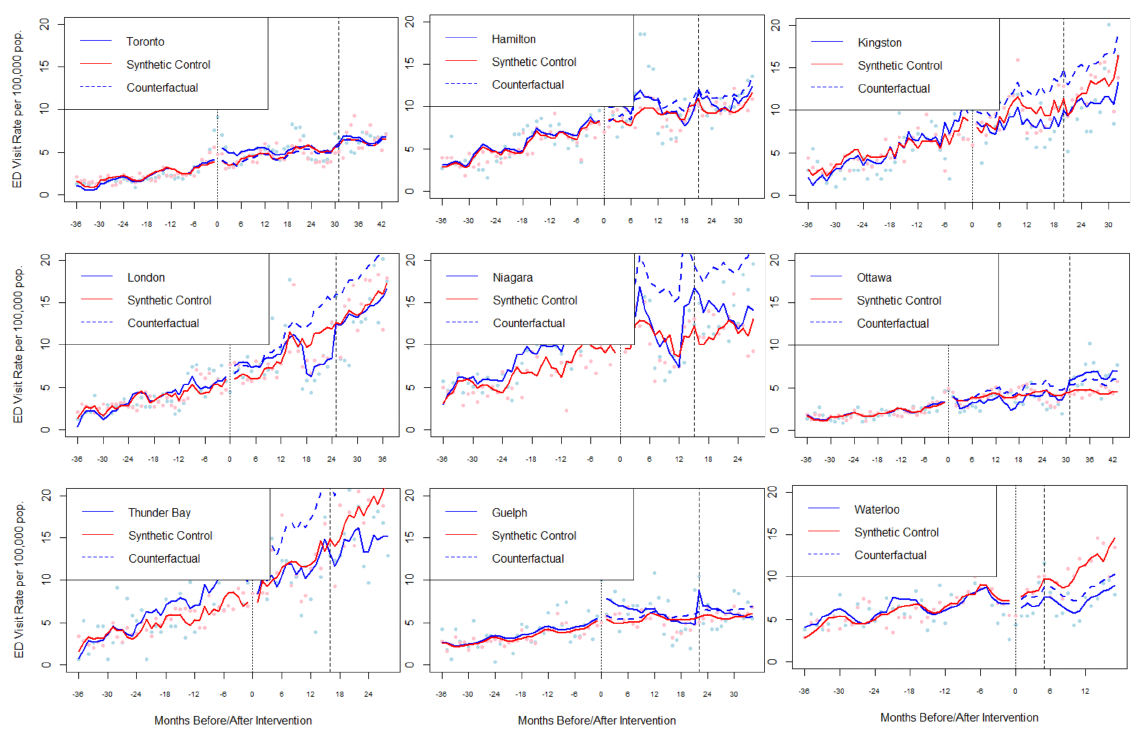

Figure 1. Controlled interrupted time-series of opioid-related ED visits plotted per treated public health unit – synthetic control pair, with counterfactual; Dark vertical dashed line = March 2020; Counterfactual represents the expected outcome rates were the intervention set to 0 in the regression models

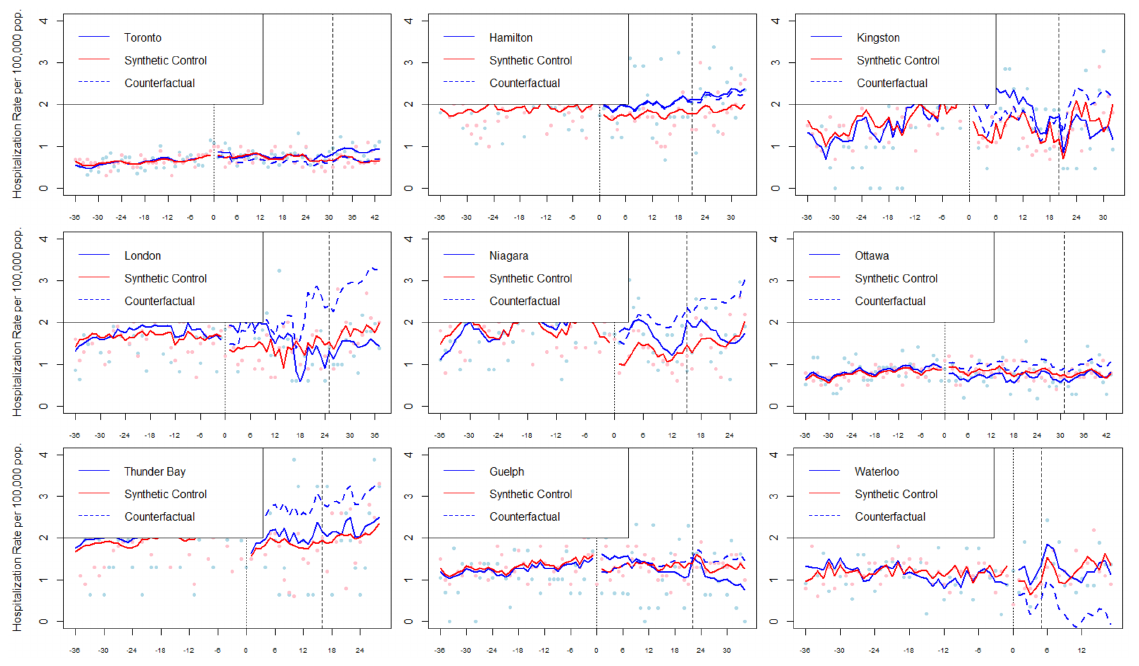

Figure 2. Controlled interrupted time-series of hospitalizations plotted per treated public health unit – synthetic control pair, with counterfactuals; Dark vertical dashed line = March 2020; Counterfactual represents the expected outcome rates were the intervention set to 0 in the regression models

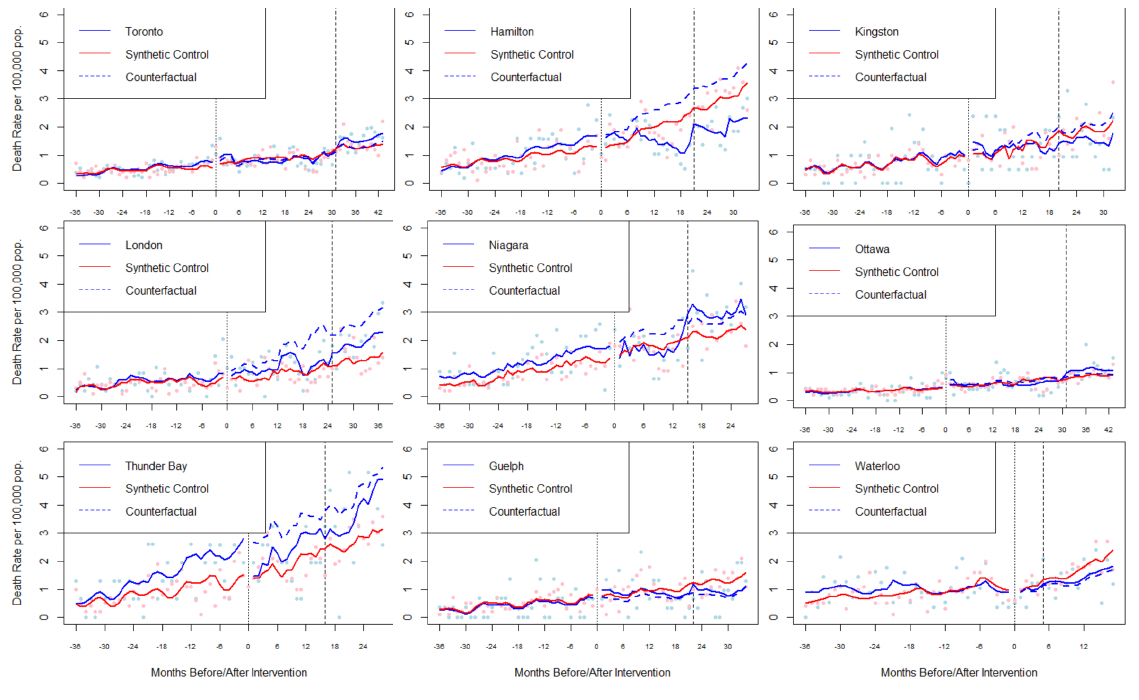

Figure 3. Controlled interrupted time-series of mortalities plotted per treated public health unit – synthetic control pair, with counterfactuals; Dark vertical dashed line = March 2020; Counterfactual represents the expected outcome rates were the intervention set to 0 in the regression models

SUPPLEMENT DATA SOURCES

1.	Medical Officer of Health, *Toronto Overdose Action Plan: Status Report 2018*, in *Report for Action*. 2018, City of Toronto: Toronto, ON.

2.	CBC News, *Toronto's first permanent supervised injection site opens downtown*, in *CBC News*. 2017: Toronto, ON.

3.	Surveillance & Epidemiology. *Toronto Overdose Information System*. 2021 16 September 2021]; Available from: <https://public.tableau.com/app/profile/tphseu/viz/TOISDashboard_Final/ParamedicResponse>

4.	Draaisma, M. and I. Lucs, *The Works superised injection site reopens with physical distancing measures in place*, in *CBC News*. 2020: Toronto, ON.

5.	Toronto Drop-In Network, *Overdose Prevention Site, Supervised Injection Services and Safe Consumption Service Hours List*. 2018: Toronto, ON.

6.	Harm Reduction TO. *Supervised Consumption Services: OPS, SIS/SCS, & CTS*. 24 May 2019 August 30 2019]; Available from: <https://harmreductionto.ca/sis-ops-scs-cts>.

7.	Toronto Public Health. *@TOPublicHealth*. 2020 16 September 2021]; Available from: <https://twitter.com/topublichealth/status/1259885006396284929>.

8.	CBC News, *Toronto opens new safe injection site at Fred Victor Centre*, in *CBC News*. 2018: Toronto, ON.

9.	Toronto Central Health Line. *Queen and Jarvis Site - Supervised Consumption Service*. 2021 1 October 2021]; Available from: <https://www.torontocentralhealthline.ca/displayservice.aspx?id=182489>.

10.	Fred Victor. *Fred Victor's response to the COVID-19 pandemic*. 2020 12 October 2021]; Available from: <https://www.fredvictor.org/fred-victors-response-to-the-covid-19-pandemic/>.

11.	Pride Toronto, *Community Resources in Response to COVID-19*. 2020.

12.	Mullin, M., *Citing overdose 'crisis' police to allow unsanctioned Toronto injection site*, in *CBC News*. 2017: Toronto, ON.

13.	Contenta, S. *Toronto Star: Overdose prevention workers save thousands of lives, but who's saving theirs?* 2019 1 September 2021]; Available from: <https://www.srchc.ca/news/tag/ops/>.

14.	CTVNews.ca Staff, *Unsanctioned pop-up safe-injection site opens in Toronto*, in *CTV News*. 2017: Toronto, ON.

15.	News Staff, *Safe injection site opens in Parkdale*, in *CityNews*. 2018: Toronto, ON.

16.	Parkdale Queen West Community Health Centre. *Hours, Location & Phone Numbers*. 2018 12 September 2021]; Available from: <https://pqwchc.org/supervised-injection-service-open-house/>

17.	City of Toronto. *Supervised Consumption Services*. 2021 16 September 2021]; Available from: <https://www.toronto.ca/community-people/health-wellness-care/health-programs-advice/supervised-injection-services/>.

18.	Parkdale Queen West Community Health Centre. *Supervised Consumption Services (SCS) - Queen West Site*. 2020 5 September 2021]; Available from: <https://pqwchc.org/programs-services/harm-reduction/supervised-consumption-services-scs/>.

19.	Parkdale Queen West Community Health Centre, *Frequently Asked Questions (FAQ) about the Overdose Prevention Site at Parkdale*. 2018, Parkdale Queen West Community Health Centre: Toronto, ON.

20.	Toronto Central Health Line. *Parkdale Queen West Community Health Centre - Parkdale Site*. 2021 1 October 2021]; Available from: <https://www.torontocentralhealthline.ca/displayservice.aspx?id=167074>.

21.	Jones, R.P., *New temporary safe injection sites open in Kensington, Regent Park*, in *CBC News*. 2018: Toronto, ON.

22.	Regent Park Community Health Centre, *OPS Pamphlet*, R.P.C.H. Centre, Editor. 2020: Toronto, ON.

23.	Kolla, G., R. Penn, and C. Long, *Evaluation of the Overdose Prevention Sites at Street Health and St. Stephen's Community House*. 2019, Street Health and St. Stephen's Community House: Toronto, ON.

24.	Street Health Community Health Centre, *Street Health Overdose Prevention Site*, S. Health, Editor. 2018.

25.	Toronto Drop-In Network. *Evaluation of the Overdose Prevention Services at St. Stephen's Community House and Street Health*. 2020 16 September 2021]; Available from: <https://www.tdin.ca/resource.php?id=671>.

26.	St. Stephen's Community House. *Temporary overdose prevention site at 260 Augusta Ave.* 2018; Available from: <https://www.sschto.ca/About-Us/In-The-News/News/Overdose-Prevention-Site-at-260-Augusta-Ave>.

27.	City of Hamilton. *Consumption and Treatment Services (CTS)*. 2018 16 September 2021]; Available from: <https://www.hamilton.ca/city-initiatives/strategies-actions/consumption-and-treatment-services#:~:text=June%205%2C%202018%20Urban%20Core,Centre%20and%20Shelter%20Health%20Network>.

28.	Hamilton Urban Core Community Health Centre. *Consumption & Treatment Service*. 2018 [cited 2021; Available from:

<http://hucchc.com/health-and-wellness-programs/harm-reduction/>.

29.	Hamilton Public Library. *Overdose Prevention Site*. 2020 18 September 2021]; Available from: <https://redbook.hpl.ca/node/14141>.

30.	Hurdle, L., *Consumption and Treatment Services - Street Health Centre*. 2019, City of Kingston: Kingston, ON.

31.	Ferguson, E., *Ministry to set up overdose prevention site*, in *The Kingston Whig Standard*. 2018: Kingston, ON.

32.	Kingston Frontenac Lennox and Addincton Public Health. *Consumption and Treatment Services*. 2020 16 September 2021]; Available from: <https://www.kflaph.ca/en/healthy-living/consumption-and-treatment-services-site.aspx>.

33.	Middlesex-London Health Unit. *Temporary Overdose Prevention Site (TOPS)*. 2018 16 September 2021]; Available from: <https://www.healthunit.com/temporary-overdose-prevention-site>.

34.	CBC News, *Take a tour of London's Temporary Overdose Prevention Site*, in *CBC News*. 2018: Toronto, ON.

35.	Kitching, S., *Overdose Prevention Site to Open Monday*. 2018, Blackburn News: London, ON.

36.	Lupton, A. *COVID-19 puts new focus on space constraints at London's drug-use site*. 2020 16 September 2021]; Available from: <https://www.cbc.ca/news/canada/london/covid-19-puts-new-focus-on-space-constraints-at-london-s-drug-use-site-1.5590576>.

37.	Clementson, L. *St. Catharines home to first supervised consumption site in Niagara region*. 2019 16 September 2021]; Available from: <https://www.cbc.ca/news/canada/hamilton/st-catharins-consumption-sit-1.4963875>.

38.	The Welland Tribune, *Positive living Niagara facing challenges during pandemic*, in *Niagara Falls Review*. 2020: Niagara Region.

39.	Ottawa Prevention, *Overdose Prevention Ottawa Summary Report*. 2018: Ottawa, ON.

40.	Raymond, T., *Ottawa Public Health to expand supervised consumption services at Clarence Street*, in *CTV News*. 2018: Ottawa, ON.

41.	CBC News, *Injection site scaling back hours due to funding shortfall*, in *CBC News*. 2019: Ottawa, ON.

42.	Crawford, B., *City's first permanent supervised injection site opens at Sandy Hill Community Health Centre*, in *Ottawa Citizen*. 2018: Ottawa, ON.

43.	Jones, R.P., *Injection sites scales back physical distancing rules to prevent ODs*, in *CBC News*. 2020: Ottawa, ON.

44.	DelVillano, S., et al., *At-a-glance - Supervised Injection Services: a community-based response to the opioid crisis in the City of Ottawa, Canada.* Health promotion and chronic disease prevention in Canada : research, policy and practice, 2019. **39**(3): p. 112-115.

45.	Payne, E., *Province supports new Inner City Health supervised injection site*, in *Ottawa Citizen*. 2017: Ottawa, ON.

46.	Fagan, L., *ByWard Market supervised injeciton trailer to get permanent home*, in *CBC News*. 2019: Ottawa, ON.

47.	Reynolds, C., *Supervised injection sites across Canada hampered by COVID-19*, in *The Canadian Press*. 2021.

48.	Whan, C., *New temporary overdose prevention site opens in Somerset West*, in *Global News*. 2018: Ottawa, ON.

49.	Raymond, T., *Somerset West CHC launches new supervised drug consumption site*, in *CTV News*. 2018: Ottawa, ON.

50.	Pivot Legal Society. *Canada's supervised consumption and overdose prevention sites*. 2020 23 April 2021]; Available from: <https://www.pivotlegal.org/scs_ops_map>.

51.	Pivot Legal Society. *Canada's Supervised Consumption and Overdose Prevention Sites*. 2019 16 September 2021]; Available from: <https://www.pivotlegal.org/scs_ops_map>.

52.	Diaczuk, D., *Overdose prevention site now open*, in *TBnewswatch*. 2018: Thunder Bay, ON.

53.	NorWest Community Health Centres. *NorWest CHC is expanding its harm reduction services*. 24 March 2020 16 September 2021]; Available from: <https://www.norwestchc.org/locations/thunder-bay/programs/path-525>.

54.	211 Ontario North. *Norwest community health centres: Path 525*. 2020 16 September 2021]; Available from: <https://211north.ca/record/65782603/>.

55.	Groleau, C., *Overdose prevention site in Guelph logs 430 visits since opening*, in *CBC News*. 2018: Kitchener-Waterloo, ON.

56.	Wellington-Duffering-Guelph, P.H. *Where to get take-home naloxone kits*. 2018 16 September 2021]; Available from: <https://www.wdgpublichealth.ca/your-health/opioids-and-naloxone/local-opioid-resources>.

57.	Armstrong, K., *Guelph safe injection site to double its capacity*, in *GuelphToday*. 2020: Guelph, ON.

58.	Centre, G.C.H. *Reduced Health Services Hours*. 2020 16 September 2021]; Available from: <https://guelphchc.ca/reduced-health-service-hours/>.

59.	CBC News, *Here's what Kitchener's consumption treatment site looks like*, in *CBC News*. 2019: Kitchener-Waterloo.

60.	Senoran, H., *Permanent CTS site coming to downtown Kitchener one year after temporary site opened*, in *CTV News*. 2020: Kitchener, ON.

61.	The Corporation of the City of Sault Ste. Marie, *Council Correspondence*. 2019, City of Sault Ste. Marie: Sault Ste. Maria, ON.

62.	Kelly, B., *Overdose does not discriminate*, in *The Brantford Expositor*. 2019: Sault Ste. Marie, ON.

63.	The Sault Star, *News briefs*, in *The Sault Star*. 2019: Sault Ste. Marie.

64.	Brant County Health Unit, *Annual Report 2017*. 2017.

65.	Brant County Board of Health, *Regular Agenda*, in *Opioid Surveillance*. 2018: Brant County, ON.

66.	Brant County Health Unit. *Brant/Brantford Opioid Information System*. 2021 16 September 2021]; Available from: <https://www.bchu.org/StatsAndReports/Pages/Opioid-Information.aspx>.

67.	Chatham - Kent Public Health, *Annual Report 2017*. 2017: Chatham, ON.

68.	Municipality of Chatham-Kent, *Improvements for CK Public Health's Harm Reduction Program*, in *Community Human Services - Public Health Unit*. 2021: Chatham, ON.

69.	Chatham - Kent Public Health, *Opioid Surveillance Summary*. 2021.

70.	City of Hamilton. *Hamilton Opioid Information System - Naloxone*. 2021 1 October 2021]; Available from: <https://www.hamilton.ca/public-health/reporting/hamilton-opioid-information-system-naloxone>.

71.	Paddon, N., *Public health handing out more naloxone kits*, in *The Hamilton Spectator*. 2018: Hamilton, ON.

72.	Tadrous, M., et al., *Naloxone Distribution Across Ontario*. 2019, Ontario Drug Policy Research Network: Toronto, ON.

73.	Public Health Grey Bruce, *Grey Bruce Opioid Response Plan*. 2020: Grey Bruce, ON.

74.	Public Health Grey Bruce, *Opioid Situation Report #1*. 2021: Grey Bruce, ON.

75.	Health and Social Services Haldimand and Norfolk, *Health Unit Opioid Management Plan*. 2018, Haldimand Norfolk Health and Social Services: Haldimand, ON.

76.	Haliburton, K., Pine Ridge District Health Unit, *2019 Annual Report: Strong Roots in our Communities*. 2020: Ontario.

77.	Halton Region. *Opioid Reporting*. 2021 September 2021 1 October 2021]; Available from: <https://www.halton.ca/For-Residents/Public-Health/Health-Statistics/Opioid-Reporting>.

78.	Hastings Prince Edward County Public Health, *The impact of opioids and other drugs in Hastings and Prince Edward Counties*. 2019, Hastings Prince Edward Public Health: Belleville, ON.

79.	Hastings Prince Edward County Public Health, *2019 Annual Report*. 2021, Hastings Prince Edward Public Health: Belleville, ON.

80.	Kingston Community Health Centres, *Annual Report 2017/18 Moving Upstream: Addressing the Social Determinants of Health*. 2019: Kingston, ON.

81.	Lambton Public Health, *2018 Annual Report*. 2019: Lambton, ON.

82.	Ontario Association of Fire Chiefs. *Analysis: Deep in the opioid crisis in Southwestern Ontario*. 2020; Available from: <https://www.oafc.on.ca/article/analysis-deep-opioid-crisis-southwestern-ontario>.

83.	Middlesex-London Health Unit. *Opioids - What is the situation?* 2021 16 September 2021]; Available from: <https://www.healthunit.com/opioids-middlesex-london#naloxone>.

84.	Niagara Region. *Naloxone use and distribution in Niagara (2019 - 2021)*. 2021 1 November 2021]; Available from: <https://www.niagararegion.ca/living/health_wellness/alc-sub-abuse/drugs/opioids.aspx#topic5>.

85.	CBC News, *Naloxone use on the rise in Thunder Bay, health unit says*, in *CBC News*. 2019: Thunder Bay, ON.

86.	Northwestern Health Unit, *2017 Public Health Report Card*. 2020, Northwestern Public Health.

87.	Northwestern Health Unit, *Moving Forward: 2018 Public Health Report Card*. 2020, Northwestern Public Health.

88.	Ottawa Board of Health, *Subject: Next steps in harm reduction and overdose prevention*. 2019, Ottawa Public Health: Ottawa, ON.

89.	Ottawa Public Health, *2017 Annual Report*. 2018, Ottawa Public Health: Ottawa, ON.

90.	Ottawa Public Health, *Subject: Ottawa overdose prevention and response task force: Update*. 2017, Ottawa Public Health: Ottawa, ON.

91.	Region of Peel, *Peel Opioid Strategy: A local response*. 2019.

92.	Porcupine health unit, *2017 Annual Report*. 2018.

93.	Porcupine health unit, *Medical officer of health report to the board of health*. 2020, Porcupine Public Health.

94.	Renfrew County and District Health Unit, *2017 Annual Report*. 2018.

95.	Renfrew County and District Health Unit, *2018 Annual Report*. 2019.

96.	Renfrew County and District Health Unit, *2019 Annual Report*. 2020.

97.	Simcoe Muskoka District Health Unit, *2018 Status Update*, in *Simcoe Muskoka Opioid Strategy*. 2019.

98.	Simcoe Muskoka District Health Unit, *Simcoe Muskoka Opioid Strategy Scorecard*. 2020.

99.	Public Health Sudbury District, *A study to explore the need for and feasibility of implementing supervised consumption services in the City of Greater Sudbury*. 2021, Public Health Sudbury District.

100.	Public Health Sudbury District. *Naloxone Distribution*. 2021 11 November 2021]; Available from: <https://www.phsd.ca/health-topics-programs/alcohol-drugs/community-drug-strategy/research/opioid-surveillance/naloxone/>.

101.	Thunder Bay District Health Unit. *Opioid Information System*. 2021 2 November 2021]; Available from: <https://www.phsd.ca/health-topics-programs/alcohol-drugs/community-drug-strategy/research/opioid-surveillance/naloxone/>.

102.	Medical Officer of Health, *Toronto Overdose Action Plan: Status Report 2019*, in *Report for Action*. 2019, City of Toronto: Toronto, ON.

103.	Medical Officer of Health, *Toronto Overdose Action Plan: Status Report 2020*, in *Report for Action*. 2020, City of Toronto: Toronto, ON.

104.	Medical Officer of Health, *Toronto Overdose Action Plan: Status Report 2021*, in *Report for Action*. 2021, City of Toronto: Toronto, ON.

105.	Region of Waterloo, *Waterloo Region Supervised Injection Services Feasibility Study*. 2018: Waterloo, ON.

106.	Regional Municipality of Waterloo, *Harm Reduction Planning, Programs and Services Update*. 2019, Region of Waterloo: Waterloo, ON.

107.	Wellington-Dufferin-Guelph, P.H., *Naloxone Distribution Program*. 2019.

108.	Windsor-Essex County Health Unit. *Opioid Overdose Statistics*. 2021 1 November 2021]; Available from: <https://www.wechu.org/reports/opioid-overdose-statistics>.

109.	Regional Municipality of York, *York Region Public Health's Opioid Action Plan*. 2019.
